# Projecting COVID-19 intensive care admissions in the Netherlands for policy advice: February 2020 to January 2021

**DOI:** 10.1101/2023.06.30.23291989

**Authors:** Don Klinkenberg, Jantien A. Backer, Nicolette F. de Keizer, Jacco Wallinga

**Affiliations:** National Institute for Public Health and the Environment, Bilthoven, The Netherlands; National Intensive Care Evaluation (NICE) Foundation, Amsterdam, The Netherlands; Leiden University Medical Centre, Leiden, The Netherlands

## Abstract

0.

**Introduction:** Model projections of COVID-19 incidence into the future help policy makers about decisions to implement or lift control measures. During 2020, policy makers in the Netherlands were informed on a weekly basis with short-term projections of COVID-19 intensive care unit (ICU) admissions. Here we present the model and the procedure by which it was updated.

**Methods:** the projections were produced using an age-structured transmission model. A consistent, incremental update procedure that integrated all new surveillance and hospital data was conducted weekly. First, up-to-date estimates for most parameter values were obtained through re-analysis of all data sources. Then, estimates were made for changes in the age-specific contact rates in response to policy changes. Finally, a piecewise constant transmission rate was estimated by fitting the model to reported daily ICU admissions, with a change point analysis guided by Akaike’s Information Criterion.

**Results:** The model and update procedure allowed us to make mostly accurate weekly projections, accounting for recent and future policy changes, and to adapt the estimated effectiveness of the policy changes based only on the natural accumulation of incoming data.

**Discussion:** The model incorporates basic epidemiological principles and most model parameters were estimated per data source. Therefore, it had potential to be adapted to a more complex epidemiological situation, as it would develop after 2020.

## 1. Introduction

Since early 2020, the COVID-19 pandemic has had a severe impact on society. To mitigate the primary impact due to infection with the SARS-CoV-2 virus, including the large burden on health care systems affecting both quantity and quality of care beyond COVID-19, public health control measures were taken that seriously affected everyday life, economically as well as socially (1, 2). A great responsibility lay with the policy-makers who had to weigh these factors into a balanced control policy, despite all uncertainties about both the positive and negative effects of control measures.

An important source of information guiding the decision to implement or lift a control measure, was the input from epidemic models. In the Netherlands, the National Institute for Public Health and the Environment (RIVM) was responsible for modelling to inform policy. Results were presented weekly to the Outbreak Management Team, the official medical advisory body to the government. Models were developed for various objectives, such as to advise on the introduction or optimisation of particular control measures, including contact tracing and vaccination (3, 4), and to assess the current state of the epidemic, by making nowcasts and estimating the effective reproduction number (5, 6).

To assess the effect of control measures in place and to allow timely decisions regarding modifications of control, there was a need for short-term projections (up to three weeks) of admissions to Intensive Care Units (ICU). Many forecasting models have been used worldwide, which make use of mechanistic transmission models or statistical techniques such as time series analyses, filtering methods, and machine learning (5, 7–14). In the Netherlands, the National Centre for Patient Exchange (LCPS), the body responsible for hospital capacity planning and patient relocations, produced weekly short-term forecasts for hospitals. These were based on extrapolation of the current growth rate (15). The Netherlands Organisation for Applied Scientific Research (TNO) developed a transmission model that was briefly used for forecasts (16), using publicly available data on ICU admissions in an ensemble smoother fitting algorithm. Both the LCPS and TNO models were not designed to project the effect of recent changes in control measures.

To provide policy-makers with short-term projections of new admissions, resulting healthcare demand and the effects of control measures, we developed a dynamic transmission model for the spread of SARS-CoV-2 in the Dutch population. The model was primarily used for projections of hospital and ICU admissions on a three-week time horizon, and a modelling procedure was designed to minimise the need for arbitrary decisions about the impact of control measures. Here we present the model, and the procedure for integration of the most recent data and recent changes in control measures. Although the procedure was followed for the entire two years with major contact-restricting control policy in place, in this article we present the model used between 28 March 2020 and 6 January 2021, which avoids the complexities of incorporating vaccination and the rise of new variants in the model description.

## 2. Methods

The model incrementally combined incoming data with a mechanistic description of recent changes in control measures, to make projections for the course of the epidemic on a three-week time horizon. On the day of analysis, the most recent data were collected to parameterize and fit the model.

### 2.1. Data

#### CBS: Population

We used the population size and age structure of the Netherlands in 2020, published by Statistics Netherlands (CBS), in 10-year age groups 0-9, 10-19, …, 70-79, 80+ years (www.opendata.cbs.nl).

#### Pienter: Contacts

In 2016-2017, the third Pienter study was conducted in the Netherlands, a nationwide seroprevalence study including a contact survey (17, 18). In this contact survey, participants aged 0-89 years (or the parents in case of child participants) filled out a contact diary over one day, in which they registered all their contacts specified by age and setting: work, school, home, leisure, transport, or other.

#### CoMix: Contacts

In 2020 an international collaboration started collecting contact survey data in several European countries (19). We used the data of the first survey conducted in the Netherlands in early April 2020 (20).

#### PiCo: Serology

In April 2020, the first serological survey in the general population for antibodies against SARS-CoV-2 was carried out in the Netherlands, followed by a second survey in June 2020 (21, 22). These surveys provided estimated seroprevalences per age group, for the mean days of sample collection of 6 April and 15 June.

#### Osiris: Case notifications

Positive SARS-CoV-2 tests are notifiable and all cases were registered in the Osiris database (23). Relevant variables for this study were: age, day of symptom onset, day of hospital admission, and potential infector. Following initial notification, records were not always updated, resulting in incomplete hospitalisation records in this database, especially after June 2020, when community-wide testing became available for everyone with COVID-19-like symptoms.

#### NICE: Hospital and ICU admissions

The National Intensive Care Evaluation registered all COVID-19 hospital admissions in the Netherlands (24). Records consisted of dates of admission and discharge or death for every patient in every ward, with pseudonymized patient identifiers allowing the linkage of potential multiple records for a single patient. As near real-time data were used, the duration of stay was censored for many patients, especially during periods of high occupancy.

#### Ownership and privacy

Most datasets contained personal information and were therefore privacy sensitive, only CBS is open data. Pienter, CoMix, and PiCo were collected and owned by RIVM. Osiris were collected by the Dutch municipal health services and reported to RIVM. NICE data from the hospitals was processed by NICE Research and Support under supervision of the NICE foundation. RIVM was permitted to use these data sources for research purposes. Anonymized case data and aggregated data were made available for public use on the RIVM website (https://data.rivm.nl/covid-19/). For the weekly data analysis the most recent individual-based data were always used.

### 2.2. Model

We used a deterministic age-structured SEEIIR compartmental model for transmission of SARS-CoV-2 in the Netherlands (Figure 1a), described with ordinary differential equations (ODEs), for nine age groups (0-9, 10-19, …, 80+ years). In this model structure, an individual starts in the *S* (susceptible) compartment, enters the *E*^1^ (latently infected) compartment by infection at rate *λ*(*t*), and then moves through each of the *E*^2^, *I*^1^, and *I*^2^ compartments into *R* (immune or dead), with each transition made at the same rate *γ*. Cases were not infectious in the *E*^1^, *E*^2^, and *R* compartments, and were equally infectious in both *I* compartments. This structure served to obtain the desired generation interval distribution (Figure 1b). The model was initialized on 12 February 2020 with *y*_0_/4 individuals in each age group in each of the four compartments *E*^1^, *E*^2^, *I*^1^, and *I*^2^. The infection incidence *y_i_*(*t*) in age group *i*, i.e. the daily rate of new infections caused by susceptibles making contacts with infectives in both infectious classes (*I* = *I*^1^ + *I*^2^), was modelled as

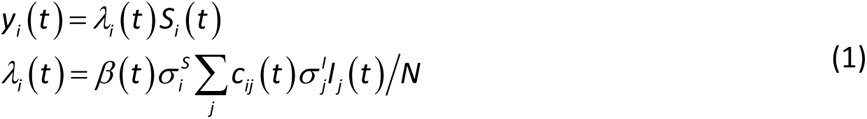

**Figure 1.**
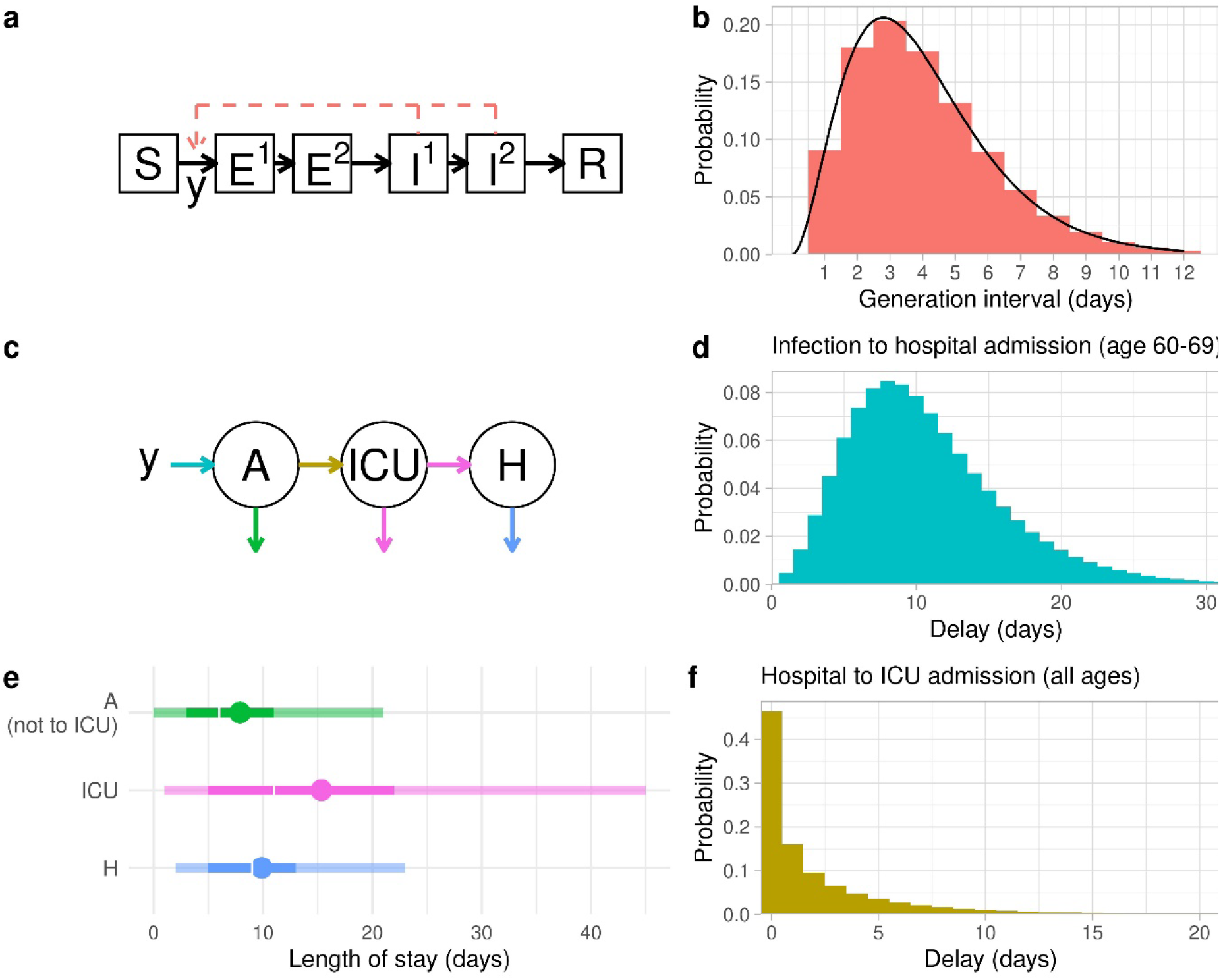
Schematic of the model, which is age-structured into nine groups 0-9, 10-19, …, 80+; (a) the dynamic SEEIIR compartmental transmission model, with incidence of infection y from S (susceptible) to E1 (exposed – first stage), and equal fixed rates between all compartments from E1 to E2 (exposed – second stage), to I1 and I2 (both infected and infectious), and finally to R (either immune or dead). Incidence y results from contacts of susceptibles (S) with infectives (I1 and I2); (b) the continuous generation interval distribution resulting from the SEEIIR structure of the transmission model (line), and the discretized version g(τ) used for the discrete-time model (histogram); (c) the clinical progression model, from infection incidence to hospital admission (general ward), transfer to the ICU, and transfer back to a general hospital ward, with outflow because of discharge or death; (d) delay distribution from infection to hospital admission (general ward); (e) length-of-stay distributions on the general ward (patients not going to ICU), on the ICU, and on the general ward (after discharge from the ICU). Points are means, white gaps are medians, solid colours are 50% range, transparant colours are 90% ranges (f), delay distribution from hospital admission to ICU admission; (d-f) situation on 6 January 2021, with colours matching the arrows in (c)

This assumes homogeneous mixing within age groups. The time-varying force of infection *λ_i_*(*t*) is the fundamental descriptor of the transmission model, and combines the underlying components infectivity, susceptibility and contact rates between individuals of all age classes, as well as how these contact rates changed due to control measures, behaviour or by other means (see next section for parameter estimation). The components of *λ_i_*(*t*) consist of:

- the population size *N*, with proportion of the population *x_i_* in age group *i* to determine the initial values *S_i_*(0) = *x_i_ N* – *y*_0_.
- the number of infected people in age group *j*, *I_j_*.
- the relative susceptibility of age group *i*, 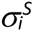, and relative infectivity of age group *j*, 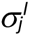.
- the rate by which each individual in age group *i* makes contacts with individuals of age group *j* (if all individuals would be of type *j*), *c_ij_*(*t*). This rate is an element of the contact matrix **C**(*t*), and is stepwise constant, depending on control measures in periods of the epidemic *T* ending at transition times *u_T_* (*u*_0_ is before the pandemic). Given the contact matrices **C***_T_*, and the transition times *u_T_*,

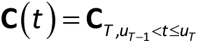
- the transmissibility parameter, *β*(*t*). This rate is stepwise constant with change points at some of the transition times *u_T_*, which reflect changes in transmissibility that are not covered by changes in the contact matrices:

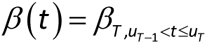

The basic reproduction number *R*_0_ of this model, defined as the mean number of secondary cases per primary case in a susceptible population, is equal to the largest eigenvalue of the next-generation matrix (25)

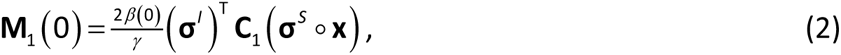

in which ◦ indicates element-wise multiplication. The reduction in contact rates due to control measures results in a relative transmission rate *φ_T_*(*t*), which is determined by replacing *β*(0) and **C**_1_ by *β*(*t*) and **C**(*t*), and calculating the ratio of largest eigenvalues

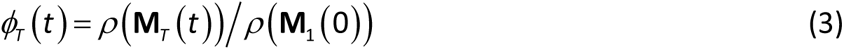

To decrease simulation time, as of 25 November 2020 this continuous-time version of the model was not used anymore and replaced (for the entire epidemic) by a discrete-time version with time steps of one day, with *I_j_*(*t*) described in terms of earlier incidence up to 12 days previously (renewal equation (25)), and generation interval distribution *g*(*τ*) (Figure 1b):

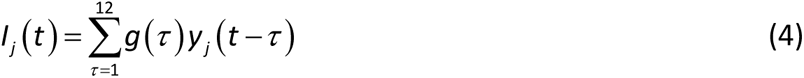

This model was initialized with incidence *y*_0_/12 in each age group on each day from 1-12 February 2020.

To simulate the daily numbers of hospital and ICU admissions and the daily numbers of discharges and deaths, the daily infection incidence *y_i_*(*t*) was used as input for the clinical progression model (Figure 1c). In this model, expected numbers of hospital and ICU admissions and occupancy are calculated, by using age-specific time-varying probabilities and delay distributions (Figure 1d-f). For instance, the expected number of ICU admissions at time *t* for age group *i* was:

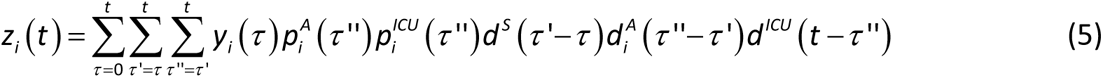

This is a convolution of three delay distributions: the delay from infection to symptom onset, the delay from symptom onset to hospital admission, and the delay from hospital admission to ICU admission. Here, 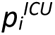(·) and 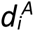(·) are the respective probabilities of hospital admission and transfer to the ICU, stratified by age and as a function of date of hospitalisation; and *d^S^*(·), 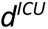(·), and *d^ICU^*(·) are the three delay distributions.

**Table 1.** Overview of model parameters, as estimated on 6 January 2021.

| Parameter | Data source <sup>a</sup> | <u>Age group</u> |  |  |  |  |  |  |  |  |
| --- | --- | --- | --- | --- | --- | --- | --- | --- | --- | --- |
|  |  | 0-9 | 10-19 | 20-29 | 30-39 | 40-49 | 50-59 | 60-69 | 70-79 | 80+ |
| Population size | CBS | 17.3 million |  |  |  |  |  |  |  |  |
| Age distribution | CBS | 0.103 | 0.116 | 0.127 | 0.122 | 0.131 | 0.145 | 0.121 | 0.088 | 0.046 |
| Incubation period distribution | (26, 27) | Weibull (shape = 2.1, mean = 5.0) |  |  |  |  |  |  |  |  |
| Generation interval distribution | (28, 29) | Figure 1b |  |  |  |  |  |  |  |  |
| Susceptibility and Infectivity | PiCo and Pienter | 1<sup>b</sup> | 3.05 | 5.75 | 3.54 | 3.71 | 4.36 | 5.69 | 5.32 | 7.21 |
| Probability of hospital admission after infection* | PiCo and NICE | 0.003 | 0.000 | 0.001 | 0.004 | 0.008 | 0.017 | 0.025 | 0.049 | 0.046 |
| Mean delay symptom onset to hospital admission(days)* | Osiris | 2.3 | 5.5 | 5.1 | 5.7 | 6.5 | 5.9 | 5.7 | 5.1 | 4.3 |
| Probability of transfer to ICU from a hospital ward* | NICE | 0 | 0.062 | 0.096 | 0.110 | 0.164 | 0.202 | 0.244 | 0.196 | 0.035 |
| Probability of transfer from ICU to a hospital ward* | NICE | 0.866 | 0.866 | 0.866 | 0.866 | 0.866 | 0.829 | 0.720 | 0.573 | 0.555 |
| Delay and length-of-stay distributions | NICE | Figure 1d-f |  |  |  |  |  |  |  |  |

\* Time-varying parameters, values of 6 January 2021; <sup>a</sup> Data sources as described in the main text: CBS = Statistics Netherlands, PiCo = serological survey, Pienter = contact survey, NICE = hospital data, Osiris = case notification data; <sup>b</sup> Reference group

### 2.3. Estimation of parameters and distributions

#### Step 1: parameters and distributions (excluding contact rates and transmissibility)

Each weekly model analysis started with the collection of all parameter values other than the contact matrices **C***_T_*, initial state *y*_0_ and transmissibility parameters *β_T_*. The analyses were adapted when necessary throughout the year, when additional data sources became available, or if parameter values appeared to change over time. We describe all analyses as carried out on 6 January 2021 (Supplement).

#### Step 2: the contact matrices

In the model, contact matrices **C***_T_*describe how different age groups interact with each other in consecutive periods *T* of the pandemic during which different sets of control measures were in place. Each matrix was used up to transition time *u_T_*, the time of policy change (Figure 2a and Supplement). An exception was made for the rapid sequential deployment of control measures mid-March 2020, with associated adaptations in contact behaviour. For this period we assumed the existence of two transition times, with the same contact matrix after the first and second transition.

**Figure 2.**
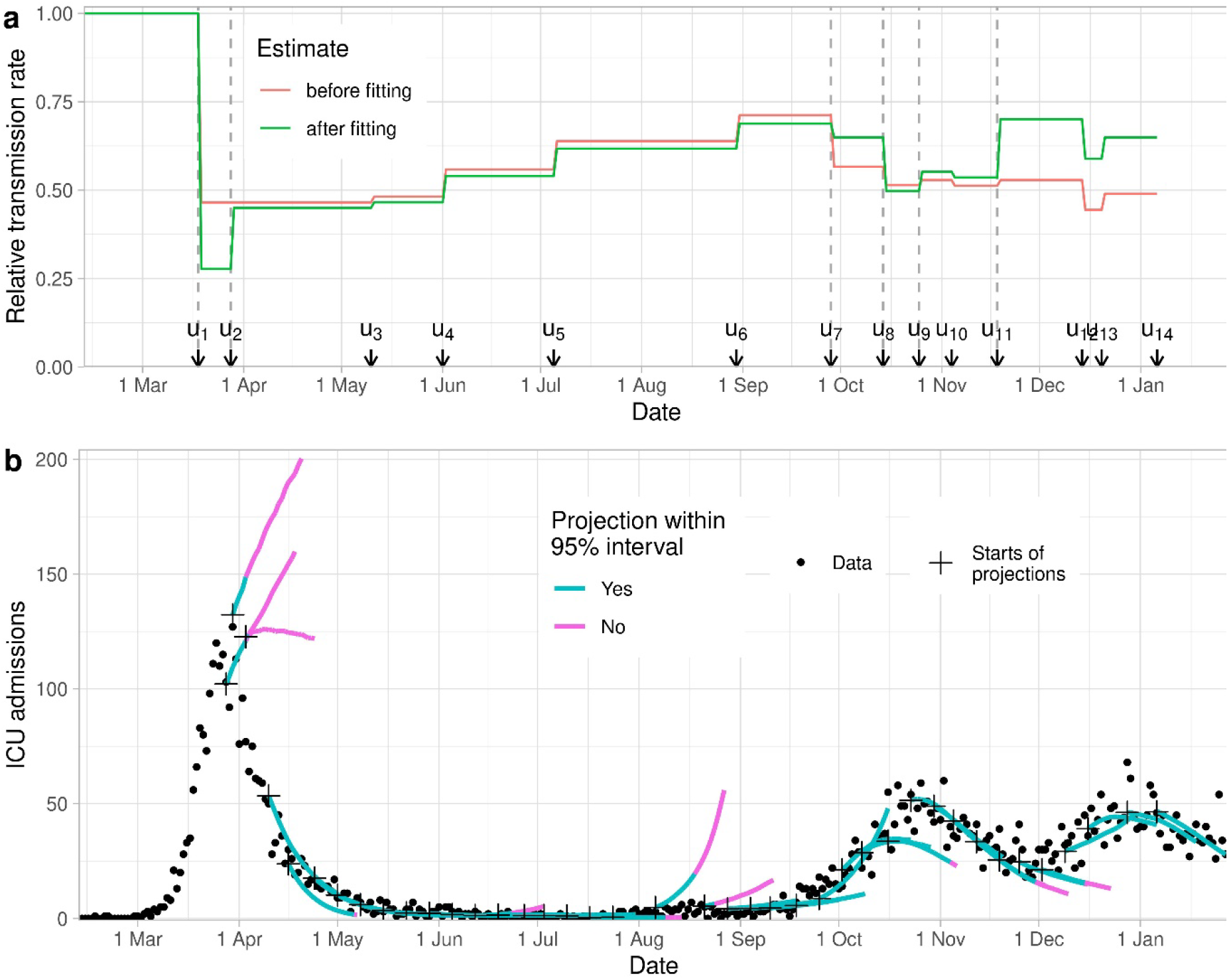
(a) Transmission rates relative to prepandemic values, before and after fitting to daily ICU admissions (analysis of 6 January 2021; Equations (2) and (3)). Arrows indicate transition times between contact matrices *u_T_*; the dotted vertical lines indicate the change points in stepwise transmissibility *β*(*t*); (b) All 40 three-week projections (starting with crosses) of numbers of ICU admissions per day until the projection made on 6 January 2021, with colour indicating when actual admissions (shown as dots) were within the 95% interval.

The dates of these transitions, *u*_1_ and *u*_2_, were estimated in step 3 (below); note that these dates do not necessarily correspond to the actual days when major policy changes were implemented.

The matrices were based on contact data collected for the Pienter study in 2017, before COVID-19 (18). In this contact survey, participants recorded their contacts over the course of a day in different settings: home, school, work, leisure, transport, and rest. COVID-19 measures can affect these contacts; how much, in which setting and in which age group will depend on the specific measure. Estimating the reduction in contacts, something that cannot be measured, was done by treating it as a Fermi problem, i.e. by breaking it down many smaller estimation problems which reduces the expected error of the problem as a whole (30, 31). For each set of COVID-19 measures, two researchers independently predicted how the measures would affect the contacts of each age group in each setting, while ensuring consistency with previous sets of measures. This yielded the Fermi estimates, i.e. consensus estimates for the contact rate reductions with an uncertainty range, which were used to resample the Pienter data and estimate the contact matrices **C***_T_* to ensure the reciprocity of the contacts between age groups (32). By uniformly sampling from the uncertainty ranges of the Fermi estimates, 200 contact matrices were obtained to describe the effect of a given set of COVID-19 control measures. On 28 May 2020, new estimates were made for all sets of control measures, to match the observations of the first CoMix contact study (19, 20), and new matrices were created.

#### Step 3: the transmissibility parameter

The transmissibility parameters *β_T_* and matrix transition times *u_T_* were required for the rate of transmission *β*(*t*). Together with the initial state *y*_0_, all *β_T_*, *u*_1_ and *u*_2_ were estimated by fitting the model to the ICU admission data, conditional on all parameter values as inferred in steps 1 and 2, and all transition times *u_T_* for *T*>2. We used the optim function in R version 3.6.0 (33) to maximize the log-likelihood, with *T*_max_ stepwise constant transmissibilities in *β*(*t*):

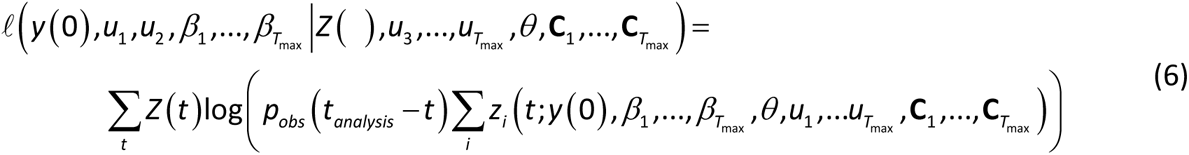

Here, *Z*(*t*) are the observed daily ICU admissions which were assumed to follow a Poisson distribution, *θ* all parameters estimated in step 1, **C***_T_* the means of all sets of 200 sampled contact matrices, *p_obs_*(*t_analysis_* - *t*) the probability of reporting at the day of analysis, and *z_i_*(*t*;…) the simulated numbers of admissions.

At each later transition time *u_T_*, the change in contact matrices from **C***_T_* to **C***_T_*_+1_ reflected the anticipated changes in contact behaviour caused by the policy change at that moment, as estimated in step 2. If the changes in behaviour were correctly reflected in the matrices, it could be assumed that *β_T_*_+1_ = *β_T_*, thus reducing the number of model parameters. As a consequence, the stepwise constant transmissibility parameter *β*(*t*) would have fewer changepoints (where *β_T_*_+1_ ≠ *β_T_*) than there are transition times. We fitted models with changepoints at different subsets of transition times *u_T_*, and selected the set with the lowest value of the Akaike’s Information Criterion (AIC, (34)). From the final selected model, point estimates were obtained with a covariance matrix for all stepwise constant transmissibilities and the initial number of infected persons, calculated from the Hessian matrix obtained with the optim function in R.

### 2.4. Model simulations: projections and scenario analyses

The parameter estimates were used to conduct 200 model simulations with different parameter sets to reflect uncertainty. Constant values were taken for the parameters estimated in step 1, and 200 samples of the parameters estimated in step 3. For the period up to two weeks prior to the analysis date, the means of the sampled contact matrices were used.

For the period after two week prior to the analysis date, additional uncertainty was added. First, all 200 samples of each contact matrix were used because contact behaviour in that period was not yet adjusted by fits to ICU data in step 3. Second, as no data were yet available to support potential future change points in transmissibility, we assumed that the transmissibility parameter remained constant, multiplied by random noise (range 0.98-1.02) to reflect the uncertainty inherent in this assumption. The simulations resulted in 200 time courses of expected ICU admissions per day. To account for stochasticity in admissions, we used each once as mean of a Poisson distribution to simulate 200 stochastic outcomes per day. Finally, mean ICU admissions and 95% intervals for each day were plotted and presented as results.

### 2.5 Model development during the pandemic

Up to 6 January 2021 the model and estimation procedure has undergone several changes, such as implementation of the SEEIIR compartments (instead of SIR), and of age-dependent susceptibility and infectivity. Also, alternatives have been explored but not implemented, such as a negative binomial likelihood and an MCMC procedure to fit the model to data, which was too time-consuming for weekly fits. Changes to the model were carefully checked in sensitivity analyses when implemented (e.g. the transition from the continuous-time to discrete-time model, see Supplement), but not systematically challenged thereafter unless evidence became available to do so.

## 3. Results

Towards the end of the first COVID-19 wave in April 2020, lengths of stay in hospital and ICU decreased and then remained constant throughout the rest of the study period (Supplement). For instance, for patients aged 60-69 years who ended up in ICU, it took on average 11 days between infection and admission to the hospital general ward (Figure 1d), then an average of two days to be transferred to the ICU (Figure 1f), where they stayed on average 15 days, with 10% of patients staying longer than 34 days (Figure 1e). After discharge from the ICU, they stayed on average a further 10 days in a general ward before being completely discharged (Figure 1e).

To assess the performance of the estimation procedure for the contact matrices and the transmissibility parameter *β*(*t*) (see Supplement for numerical results), we summarised the contact matrices by their associated reduction in transmission rates before fitting to ICU admission, *φ_T_*(0), and after fitting to ICU admissions, *φ_T_*(*t*) (Equation (3), Figure 2a). The reduction in transmission rates before fitting describes the effects of the control measures according to the estimated contact matrices. There was a large drop in transmission around 18 March, when the first lockdown started. It then gradually increased until September 2020, when more measures were imposed until reaching a new minimum just before the Christmas holiday of 2020.

The reduction in transmission rates after fitting to the ICU admissions combines the estimated transmission matrices and the stepwise constant transmissibility function *β*(*t*). In this function, a total of six changepoints were needed as of 6 January 2021. The first was placed at the start of lockdown (18 March 2020) and the second about one week later. These timepoints reflect stepwise implementation of control measures, and suggested a short-lasting extreme decrease in contact behaviour before settling to a level approximately matching the contact matrix estimate. The estimates of the exact timing and magnitude of this short decrease in March 2020 changed several times during the year when the model changed, but that did not affect projections (Supplement). No further change points were needed until the end of September, after which three were needed during October and November in which infections were rising rapidly, but with only minor changes in transmissibility. A large correction was needed mid-November at *u*_11_ (Figure 2a), when transmissibility was estimated much higher than anticipated based on the control measures only.

The resulting three-week projections during this first year of COVID-19 modelling show that most were accurate in the sense that the actual (i.e., later observed) number of ICU admissions fell within the 95% prediction intervals (Figure 2b). ICU admission projections were too high at the end of March during the first peak, and at some instances during August 2020; projections were occasionally too low in November 2020. These projections improved when new data became available, providing evidence for new changepoints (Supplement).

## 4. Discussion

During the Dutch COVID-19 epidemic, real-time short-term projections of COVID-19 ICU admissions were provided to policy makers in the Netherlands. Every week, the most recent data were incrementally incorporated into an age-structured transmission model in three steps: firstly, transition probabilities and delay distributions for the clinical progression model were estimated, as well as most parameters of the transmission model; secondly, contact matrices were estimated for each set of control measures; thirdly, a time-varying transmissibility parameter was estimated that provided the best fit to the observed daily numbers of ICU admissions. This data assimilation process avoided a combinatorial explosion of parameter inferences for a model that increased in complexity over time, while minimizing arbitrary decisions about the impact of control. The resulting model is semi-mechanistic: it combines statistical flexibility for an accurate and parsimonious description of the incidence of ICU admission in real time with a causal interpretation of the changes in contact matrices that allowed the anticipation of effects of recent and planned changes in control measures that are not yet detectable in the incoming data. The model complies with the theoretical (and practical) requirement that the model’s dynamical behaviour explains the observed dynamics, and, more importantly, the projections match the actual ICU admissions remarkably well.

A substantial change in the transmissibility parameter was estimated at only two points in the study period to correct the epidemic growth rate implied by the contact matrices (between *u*_1_ and *u*_2_, and at *u*_11_, Figure 2a). The first of these corrections was for March 2020, in the first week of the first lockdown, when the estimated transmissibility parameter was lower than anticipated for one week before increasing to the anticipated level. A similar brief drop in contact rates in this period in the Netherlands was reported by van Wees et al (35). The estimates likely reflect an actual behavioural change in the population in response to the rapidly rising number of daily hospitalizations with COVID-19. The second correction was for November 2020, during the second epidemic wave, when estimated transmissibility parameter was higher than anticipated. This may in part be explained by a decreasing adherence of the population to control measures. Another explanation is a seasonal change in transmissibility, with higher values in winter. Indeed, after implementing a seasonally dependent transmission parameter into the model (implemented in January 2021, subsequent to our study period), a much smaller correction was needed. Naturally, short-term projection intervals did not capture the actual reported numbers of ICU admissions at these two time points, with projections improving shortly thereafter, when new data became available (Figure 2b and Supplement). In addition some projection interval failed to capture admission in August, when the numbers of daily ICU admissions were low. Overall, the projections captured the later reported dynamics in ICU admissions reasonably well, which is a prequisite for informing policy-makers.

In practice, the need for timely short-term projections required making a number of assumptions, the impact of which should be considered. The transmission model used here is deterministic and assumes homogeneous mixing within age groups for the whole Dutch population. Even though the model did not capture spatial heterogeneity and local stochastic effects, the simulations captured the national average well for a population of 17 million persons. Many parameters and delay distributions were estimated independently, which risks underestimating uncertainty and ignoring potential correlation between parameter values. However, it seems that less accurate projections were generally caused by differences in anticipated contact rates – later corrected by estimates of the time-dependent transmissibility, as explained above – and not by overly narrow prediction intervals resulting from the use of independent estimates. By estimating most parameters in the first step, the inferential procedure was time-efficient, ensuring sufficient time to update all weekly estimates and to integrate the most recent data into the transmission model. This efficiency proved vital when more complex extensions of the model were deployed later in the pandemic. Estimation of the contact matrices was done by combining contact data from before the pandemic and reductions estimated by two researchers in a standardized protocol. This was feasible given the time constraints and turned out well. A major uncertainty in this protocol was future adherence to control measures. This may be inferred from behavioural surveys, but more research is needed to evaluate this possibility for real-time prediction.

This study has focused on the epidemic dynamics over the period February 2020 to January 2021 a period when the vast majority of the population was susceptible to infection and recurrent waves were a direct consequence of contact behaviour. Because the model incorporates basic principles dictated by epidemic theory, it had potential for use in the range of epidemiological situations that occurred since then. Firstly, the model can be readily adapted to include compartments and stratified parameters for vaccinated persons such that it could be used to make projections in a situation with COVID-19 vaccination. Secondly, the model can be adapted to include dynamic transmissibility and immune escape rates, when data on genomic surveillance provide evidence thereof, caused by variants of the virus. Thirdly, the loss of immunity can be modelled to facilitate the description of endemic dynamics of an infection spreading in a population that has been vaccinated or previously infected. When the epidemiological situation becomes more complex, when more data sources are available, and when research and policy questions become more interrelated, the approach of relating the transmission model to incoming data proves efficient and flexible. We believe this testifies to its practical merits.

## Supporting information

Supplemental Methods and Results

## 5. Acknowledgements

We gratefully acknowledge the Data Analytics, Research and Automated Reporting (DARA) Team at the National Institute for Public Health and the Environment for processing all COVID-19 surveillance data, and in particular Priscila de Oliveira Bressane Lima for her help with the NICE data. We thank Eric Vos for help with the PiCo data, and Scott McDonald for editing the manuscript.

## Ethical statement

Collection of the PiCo serological data was approved by the Medical Ethics Committee MEC-U, the Netherlands (Clinical Trial Registration NTR8473). For collection of the OSIRIS and NICE surveillance data, the Centre for Clinical Expertise at the RIVM verified that it does not fulfill the specific conditions stated in article 1 of the Dutch law for Medical Research Involving Human Subjects (WMO), and that approval by an ethical research committee was therefore unnecessary (RIVM study number EPI-623).

## Data and code availability

Most data contain person-disclosing information and cannot be shared. To reproduce all analyses of the last week of this study (6 January 2021), all code is made available with synthetic data to run the data analyses resulting in approximately the same parameter estimates, and with original parameter estimates to reproduce the exact simulation results. Code and synthetic data can be found at www.github.com/rivm-syso/COVID-projectionmodel.

