## Supplemental Methods and Results for "Projecting COVID-19 intensive care admissions in the Netherlands for policy advice: February 2020 to January 2021"

### Supplementary material

This is a supplement with the article P*rojecting COVID-19 intensive care admissions in the Netherlands for policy advice: February 2020 to January 2021*, by Don Klinkenberg, Jantien Backer, Nicolette de Keizer, and Jacco Wallinga. In this supplement we describe the model structure and the equations. We present the various steps for inferring model parameters. The order of these steps matter. The presentation is organized such that at each step the values of input variables have been obtained in earlier steps. We present the model version that was used on 6 January 2021, the last day of the study period.

The code used for the analyses and simulations on 6 January 2021 is available at http://www.github.com/rivm-syso/COVID-projectionmodel. To be able to use the code for the analysis of dataset with personal information, synthetic data have been created from the original data and estimation results. These synthetic data have been created from datasets with personal information, but with real persons replaced by 'fake persons'. That has been done in such a way that the analysis of the synthetic data gives approximately the same results as analysis of the original data. If real persons seem to recognise themselves in the synthetic data, then that is pure coincidence: we do not publish personal data. Statistical properties of the synthetic data other than the results in this repository do not necessarily reflect the statistical properties of the original data. These synthetic data may not be used outside this repository.

Contents

1. The model
1.1. The transmission model
1.1.1. The ODE model
1.1.2. The next-generation matrix and reproduction number
1.1.3. The discrete-time model
1.2. The clinical progression model
1.3. The observed number of daily ICU admissions

2. Parameter estimation
2.1. Step 1: parameters and distributions (excluding contact rates and transmissibility)
2.1.1. Parameter values based on literature
2.1.2. Parameter values based on demographic data from Statistics Netherlands (CBS)
2.1.3. Parameter values based on hospitalization data (NICE)
2.1.4. Parameter values based on notification data (OSIRIS)
2.1.5. Parameter values based serological survey PiCo
2.2. Step 2: parameter estimation for age-specific contact patterns
2.3. Step 3: estimation of the transmissibility parameter

3. Supplemental analyses and results
3.1. Complete results of analysis step 3, and comparison of the ODE and discrete-time models
3.2. Weekly evolution of transmissiblity parameter estimates

1. The model

1.1. The transmission model

1.1.1. The ODE model

The transmission model simulates the daily number of infections $y_i(t)$ in each age group *i*. There are nine age groups: 0-9, 10-19, … 70-79, 80+ years old. The number of individuals in each age group *i* is denoted by $N_i$.

Early in the pandemic, the simulations were done with a system of ordinary differential equations:

$$\begin{aligned}
\frac{dS_i(t)}{dt} &= -y_i(t) \\
\frac{dE_i^1(t)}{dt} &= y_i(t) - \gamma E_i^1(t) \\
\frac{dE_i^2(t)}{dt} &= \gamma E_i^1(t) - \gamma E_i^2(t) \\
\frac{dI_i^1(t)}{dt} &= \gamma E_i^2(t) - \gamma I_i^1(t) \\
\frac{dI_i^2(t)}{dt} &= \gamma I_i^1(t) - \gamma I_i^2(t) \\
\frac{dR(t)}{dt} &= \gamma I_i^2(t)
\end{aligned} \tag{S1}$$

The model was initialized (*t* = 0 is 12 February 2020) with $E_i^1(0) = E_i^2(0) = I_i^1(0) = I_i^2(0) = y_0/4$. Here, the infection incidence $y_i(t)$ in age group $i$, i.e. the daily rate of new infections, is modelled as

$$\begin{aligned}
y_i(t) &= \lambda_i(t) S_i(t) \\
\lambda_i(t) &= \beta(t)\sigma_i^S \sum_j c_{ij}(t)\sigma_j^I I_j(t)/N
\end{aligned} \tag{S2}$$

This assumes homogeneous mixing within age groups. The time-varying force of infection $\lambda_i(t)$ is the fundamental descriptor of the transmission model, and combines the underlying components infectivity, susceptibility and contact rates between individuals of all age classes, as well as how these contact rates changed due to control measures, behaviour or by other means (see next section for parameter estimation). The components of $\lambda_i(t)$ consist of:

- the population size $N$, with proportion of the population $x_i$ in age group $i$ to determine the initial values $S_i(0) = x_i N$
- the number of infected people in age group $j$, $I_j$ (sum of compartments $I^1$ and $I^2$ in the SEEIIR model)
- the relative susceptibility of age group $i$, $\sigma_i^S$, and relative infectivity of age group $j$, $\sigma_j^I$
- the rate by which each individual in age group $i$ makes contacts with individuals of age group $j$ (if all individuals would be of type $j$), $c_{ij}(t)$. This rate is an element of the contact

matrix $\mathbf{C}(t)$, and is stepwise constant, depending on control measures in periods of the epidemic $T$. Given the contact matrices $\mathbf{C}_T$, and the transition times $u_T$, $\mathbf{C}(t)=\mathbf{C}_{T,u_{T-1}<t\le u_T}$.

- the transmissibility parameter, $\beta(t)$. This rate is stepwise constant with change points at some of the transition times $u_T$, which reflect changes in transmissibility that are not covered by changes in the contact matrices: $\beta(t)=\beta_{T,u_{T-1}<t\le u_T}$.

1.1.2. The next-generation matrix and reproduction ratio

To further analyse this model, e.g. to estimate the change in the reproduction number, the model can be described by the time-varying next-generation matrix (1), which is defined as

$$\mathbf{M}_T(t)=\tfrac{2\beta(t)}{\gamma}\left(\boldsymbol{\sigma}^I\right)^{\mathrm{T}}\mathbf{C}_T\left(\boldsymbol{\sigma}^S\circ\mathbf{x}\right), \tag{S3}$$

in which $\circ$ indicates element-wise multiplication. In this matrix, each element $m_{ij}$ is the expected number of individuals in age group *j* that is infected by each infective in age group *i*, if the population would be completely susceptible. In this equation we explicitly decouple the contact matrix $C_T$ from time *t*, so that we can evaluate changes due to the estimated contact matrices alone (by using $\beta(0)$ and any matrix $\mathbf{C}_T$), and changes due to contact matrices and transmissibility (by using $\beta(t)$ and $\mathbf{C}_T=\mathbf{C}(t)$).

The basic reproduction number $R_0$, defined as the mean number of secondary cases per primary case in a susceptible population, in the absence of control measures, is equal to the largest eigenvalue $\rho$ of this matrix at the start of the epidemic

$$\mathbf{M}_1(0)=\tfrac{2\beta(0)}{\gamma}\left(\boldsymbol{\sigma}^I\right)^{\mathrm{T}}\mathbf{C}_1\left(\boldsymbol{\sigma}^S\circ\mathbf{x}\right), \tag{S4}$$

The relative transmission rate during control measures $\phi_T(t)$ is thus determined as the ratio of the eigenvalues of these two matrices:

$$\phi_T(t)=\rho\left(\mathbf{M}_T(t)\right)/\rho\left(\mathbf{M}_1(0)\right) \tag{S5}$$

To evaluate the change in reproduction number attributable to the change in estimated contact matrix (in Figure 2 in the main text), we calculated $\phi_T(0)$.

1.1.3. The discrete-time model

To decrease simulation time, as of 25 November 2020 the continuous-time version of the model was replaced by a discrete-time version with time steps of one day, with $I_j(t)$ described in terms of earlier incidence up to 12 days previously (renewal equation (1)), and generation interval distribution $g(\tau)$ (Figure 1b in the main text):

$$I_j(t)=\sum_{\tau=1}^{12} g(\tau) y_j(t-\tau) \tag{S6}$$

Simulations with this discrete-time model start with the number of susceptible individuals in each age group on 1 February 2020 as $S_i = N_i$. Simulations are initialized with a constant daily number of infections $y_i(t)=y_0/12$ for each age group from 1 to 12 February 2020. The transmission model runs in discrete time, with day $t=0$ corresponding to 12 February 2020. At each time step *t*, the hazard rate of infection $\lambda_i(t)$ is calculated for each age group *i* as:

$$\lambda_i(t)=\beta(t)\sigma_i \sum_j c_{ij}(t)\sigma_j \sum_{\tau=1}^{12} y_j(t-\tau) g(\tau)/N. \tag{S7}$$

Here, we assume that the age-specific susceptibilities and the age-specific infectivities are identical, so $\sigma_i^S = \sigma_i^I = \sigma_i$. This assumption was made when the model was changed to include these heterogeneities, when serological data became available showing heterogeneity in attack rates. Several assumptions were considered: (i) age-specific susceptibility only, (ii) age-specific infectivity only, and (iii) both. No differences in results were seen between options (i) and (iii). We selected option (iii) because we reasoned that age-dependent severity may be associated with infectivity, and stayed with that choice, in line with our general workflow regarding model development (Section 2.5 main text).

This hazard rate of infection $\lambda_i(t)$ determines how the daily number of new infections and number of susceptibles in each age group depend on the numbers on the previous day:

$$\begin{aligned} y_i(t) &= \left(1-\exp\left(-\lambda_i(t)\right)\right) S_i(t-1) \\ S_i(t) &= S_i(t-1) - y_i(t) \end{aligned}.$$

A comparison in results between the continuous-time and discrete-time models is given in section 3.1 of this Supplement.

#### 1.2. The clinical progression model

The daily numbers of infections $y_i(t)$ are used to calculate the daily numbers of hospital admissions, hospital discharges from the general ward (non-ICU patients), ICU admissions, ICU discharges to the general ward, and hospital discharges from the general ward (ICU patients). Hospital patients that die are included in the daily number of discharges, i.e. the length-of-stay distributions do not differ between patients that die and patients that are discharged. The progression from infection to hospital admission, and possibly ICU admission, and possibly back to a general ward, is determined by time-varying probabilities:

1. $\Pr_i^A(t)$: the probability of hospital admission in age group *i*, at time *t* at which hospital admission would occur (after the incubation period and subsequent delay to hospital admission)
2. $\Pr_i^{ICU}(t)$: the probability of transfer to ICU in age group *i*, given hospital admission at time *t*

3. $\Pr_i^H(t)$: the probability of transfer back to a hospital ward from ICU, given hospital admission at time *t*

The times at which events, such as symptom onset, hospital admission, happen are determined by probability distributions for the time period between these events:

4. $d^S \sim W(\alpha^S, \mu^S)$: for the incubation period, the time from infection to symptom onset, which follows a (discretised) Weibull distribution with shape $\alpha^S$ and mean $\mu^S$
5. $d_i^A(t) \sim NB(\alpha^A, \mu_i^A(t))$: the time period from symptom onset to hospital admission follows a negative binomial distribution where the mean depends on age and time of symptom onset
6. $d^{D_A}(t) \sim NB(\alpha^{D_A}, \mu^{D_A}(t))$: the time period from hospital admission to either discharge or death without being admitted to the ICU follows a negative binomial distribution where the mean depends on the time of hospital admission
7. $d^{ICU}(t) \sim NB(\alpha^{ICU}, \mu^{ICU}(t))$: the time period from hospital admission to transfer to the ICU follows a negative binomial distribution where the mean depends on the time of hospital admission
8. $d^{D_{ICU}}(t) \sim NB(\alpha^{D_{ICU}}, \mu^{D_{ICU}}(t))$: the time period from ICU admission to discharge or death follows a negative binomial distribution where the mean depends on the time of hospital admission
9. $d^{D_H}(t) \sim NB(\alpha^{D_H}, \mu^{D_H}(t))$: the time period from transfer from ICU back to a hospital ward to discharge or death from that hospital ward follows a negative binomial distribution where the mean depends on the time of hospital admission

We compute the daily number of ICU admissions from the daily number of infections $y_i(t)$ using the probabilities and probability distributions that are described above. We denote the resulting daily number of simulated ICU admissions as $z_i(t)$:

$$z_i(t) = \sum_{\tau=0}^{t}\sum_{\tau'=\tau}^{t}\sum_{\tau''=\tau'}^{t} y_i(\tau)\Pr_i^A(\tau'')\Pr_i^{ICU}(\tau'')d^S(\tau'-\tau)d_i^A(\tau''-\tau')d^{ICU}(t-\tau'') \quad \text{(S8)}$$

Here, $\tau$ enumerates infection days, $\tau'$ enumerates days of symptom onset, and $\tau''$ enumerates days of (potential) hospital admission.

1.3. The observed number of daily ICU admissions

The ICU admissions are reported with a short delay. The probability of an ICU admission on day *t* being reported on the day of the analysis, $t_{analysis}$, is indicated as $p_{obs}(t_{analysis} - t)$. The expected number of observed ICU admissions on day *t* is $p_{obs}(t_{analysis} - t)\sum_i z_i(t)$.

The transmission model is fitted such that the actual observed daily number of ICU admissions is close this expected daily number of ICU admissions. To obtain this fit we take the initial incidence $y_0$ and transmission parameter $\beta(t)$ (in fact, the piecewise constant values $\beta_T$) that maximizes the Poisson likelihood function for the expected number of observed ICU admissions.

2. Parameter estimation

2.1. Step 1: parameters and distributions (excluding contact rates and transmissibility)

Each weekly model analysis starts with the collection of all parameter values other than the contact matrices $\mathbf{C}_T$ and transmissibility parameters $\beta_T$. The analyses are adapted when necessary throughout the year, when additional data sources become available, or if parameter values appear to change over time. Here we describe the analyses as carried out on 6 January 2021, the final analyses without variants, seasonality, and vaccination.

2.1.1. Parameter values based on literature

*Transmission model: generation interval distribution*
The generation interval distribution $g(\tau)$ resulting from the SEEIIR model with transition rate $\gamma = 0.875$ is based on early estimates from China (2, 3). The mean generation interval of 4.0 days is regularly validated against contact tracing data, and was not adapted during the first year of the pandemic.

*Clinical progression model, function 4: incubation period distribution*
The incubation period distribution $d^S(.)$ (the time infection to symptom onset) is based on literature estimates (4, 5): $\alpha_S = 2.1$ and $\mu_S = 5$.

2.1.2. Parameter values based on demographic data from Statistics Netherlands (CBS)

*Transmission model, population size and age distribution*
From CBS (Statistics Netherlands), we obtain the population parameters:
$N = 17282160$

$$\mathbf{x}^\mathrm{T} = \begin{pmatrix} 0.103 & 0.116 & 0.127 & 0.122 & 0.131 & 0.145 & 0.121 & 0.088 & 0.046 \end{pmatrix}$$

2.1.3. Parameter values based on hospitalization data (NICE)

The NICE hospital data consist of records per patient, with their current status (at the time of analysis), and all dates of reporting, admission, transfer, or discharge/death. These are used to estimate the time-varying probabilities and probability distributions for time-to-events in the clinical progression model (functions 1 (partly), 2, 3, 6, 7, 8, and 9 listed in section 1.2), and the reporting probability distribution $p_{obs}$ for ICU admissions.

*Clinical progression model: the probability of hospital admission (function 1)*
At the end of March 2020 there was a sharp decline in probability of death after being hospitalised, which might have been caused by a change in hospital admission policy. We account for this by assuming that the probability of hospital admission decreased for those who would die, modelled by a logistic curve. In each age group we assume a fixed probability of hospital admission followed by discharge (cure) $p_i^{A+C}$, and a time-varying probability of hospital admission followed by death $p_i^{A+D}(t)$, so that the total probability of hospital admission was

$$\begin{aligned}&\Pr_i^A(t) = p_i^{A+C} + p_i^{A+D}(t) = p_i^{A+C} + \Pr_i^A(t)\, p_i^{D|A}(t) \Rightarrow \\ &\Pr_i^A(t) = \frac{p_i^{A+C}}{1 - p_i^{D|A}(t)}\end{aligned} \tag{S9}$$

From the NICE hospital data, we estimate the probability of death given admission $p_i^{D|A}(t)$, whereas the parameter $p_i^{A+C}$ is estimated using the PiCo data (see below).

The probability of death is low in young patients (1.3% among all patients under 40 years of age, in 2020). Because there is very little information about young patients early in the pandemic, we assume a parametric form for the age-dependence of the probability of dying among those that are admitted early in the pandemic (the left end of the logistic curve, at $t = -\infty$ ):

$$p_i^{D|A}(-\infty) = \frac{e^{a+bi}}{1 + e^{a+bi}} \tag{S10}$$

and we assume that the probability of hospital admission did not change for patients younger than 40 years of age:

$$p_i^{D|A}(\infty) = p_i^{D|A}(-\infty), \text{for } i \le 4 \tag{S11}$$

The relative probability of hospital admission followed by death $\rho_i(t) = p_i^{A+D}(t) / p_i^{A+D}(-\infty)$, is modelled with a logistic curve, with $\rho_i(\infty)$ being the ultimate reduction, $t_m$ the inflexion point of the logistic curve, and *r* the slope of the curve at the inflexion point:

$$\rho_i(t) = \frac{p_i^{A+D}(t)}{p_i^{A+D}(-\infty)} = \frac{1 + \rho_i(\infty) e^{r(t - t_m)}}{1 + e^{r(t - t_m)}} \tag{S12}$$

Note that $\rho_i(\infty) = 1$ for age groups $i \le 4$ because of the assumption in Equation (S11). Now, the probability of dying in the hospital in age group *i* $p_i^{D|A}(t)$ can be written in terms of Equations (S10) and (S12) :

$$\begin{aligned}&(i) \text{ using Eq (S9): } p_i^{D|A}(t) = \frac{p_i^{A+D}(t)}{p_i^{A+D}(t) + p_i^{A+C}} = \frac{p_i^{A+D}(-\infty)\rho(t)}{p_i^{A+D}(-\infty)\rho(t) + p_i^{A+C}} \\ &(ii) \text{ using} (i) \text{ at } t = -\infty \text{: } p_i^{A+C} = \frac{p_i^{A+D}(-\infty)\left(1 - p_i^{D|A}(-\infty)\right)}{p_i^{D|A}(-\infty)} \\ &(iii) \text{ using } (ii) \text{ in } (i) \text{: } p_i^{D|A}(t) = p_i^{D|A}(-\infty)\rho_i(t) \Big/ \left(1 - p_i^{D|A}(-\infty)\left(1 - \rho_i(t)\right)\right)\end{aligned} \tag{S13}$$

Fitting is done by maximizing the log-likelihood function:

$$\ell\left(a, b, r, t_m, \boldsymbol{\rho}(\infty) \middle| \boldsymbol{\iota}^D, \mathbf{t}, \mathbf{i}\right) = \sum_k \iota_k^D \log\left(p_{i_k}^{D|A}(t_k)\right) + \left(1 - \iota_k^D\right) \log\left(1 - p_{i_k}^{D|A}(t_k)\right) \tag{S14}$$

in which $\iota_k^D = 1$ indicates that individual $k$ died, $\iota_k^D = 0$ indicates recovery, $t_k$ is the time of hospital admission, and $i_k$ is the age group of individual *k*. The resulting fit is shown in Figure S1.

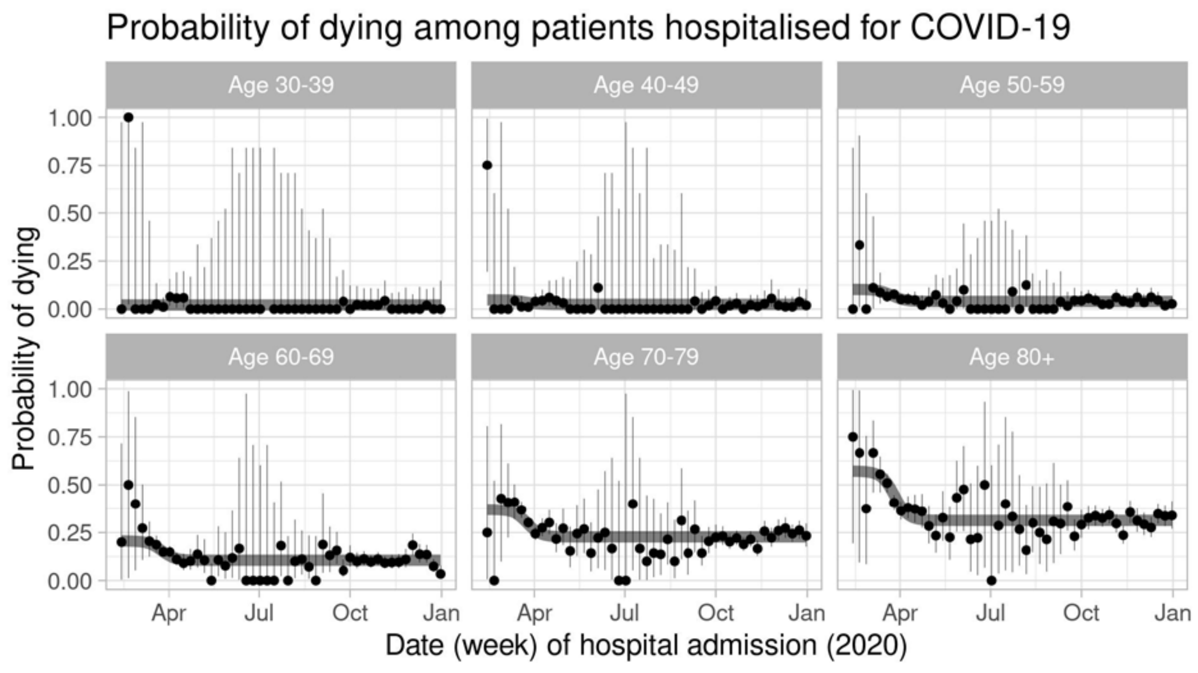


Figure S1. Estimated age-specific and time-varying probability of dying among patients hospitalised for COVID-19. The grey curves are the model fits. The points and vertical lines are weekly means and 95% confidence intervals.

Combining this result with the estimated parameters $p_i^{A+C}$ from the PiCo data (see below) gives the probability of hospital admission (Figure S10)

*Clinical progression model: the probability of transfer to the ICU (function 2)*
The probability of transfer to the ICU is modelled as a logistic curve

$$\Pr_i^{ICU}(t)=\Pr_i^{UCI}(-\infty)+\left(\Pr_i^{UCI}(\infty)-\Pr_i^{UCI}(-\infty)\right)\frac{e^{r(t-t_m)}}{1+e^{r(t-t_m)}} \quad \text{(S15)}$$

The parameters are estimated by maximizing the likelihood function given the observed outcomes for all patients admitted to the hospital

$$\ell\left(\mathbf{Pr}^{UCI}(-\infty),\mathbf{Pr}^{ICU}(\infty)\middle|r,t_m,\boldsymbol{\iota}^{ICU},\mathbf{t},\mathbf{i}\right)=\sum_k \iota_k^{ICU}\log\left(\Pr_{i_k}^{ICU}(t_k)\right)+\left(1-\iota_k^{ICU}\right)\log\left(1-\Pr_{i_k}^{ICU}(t_k)\right) \quad \text{(S16)}$$

in which $\iota_k^{ICU}=1$ indicates if individual $k$ was admitted at the ICU, $t_k$ is the time of hospital admission, and $i_k$ is the age group of individual *k*. The parameters $r$ and $t_m$ are not estimated, but fixed at the estimates obtained after maximizing Equation (S14). Patients not transferred to the ICU ( $\iota_k^{ICU}=0$ ) but still in the hospital are censored observations, as they can be transferred later, but this is ignored as the time interval from hospital admission to IC transfer was very short for most cases. The resulting fit is shown in Figure S2

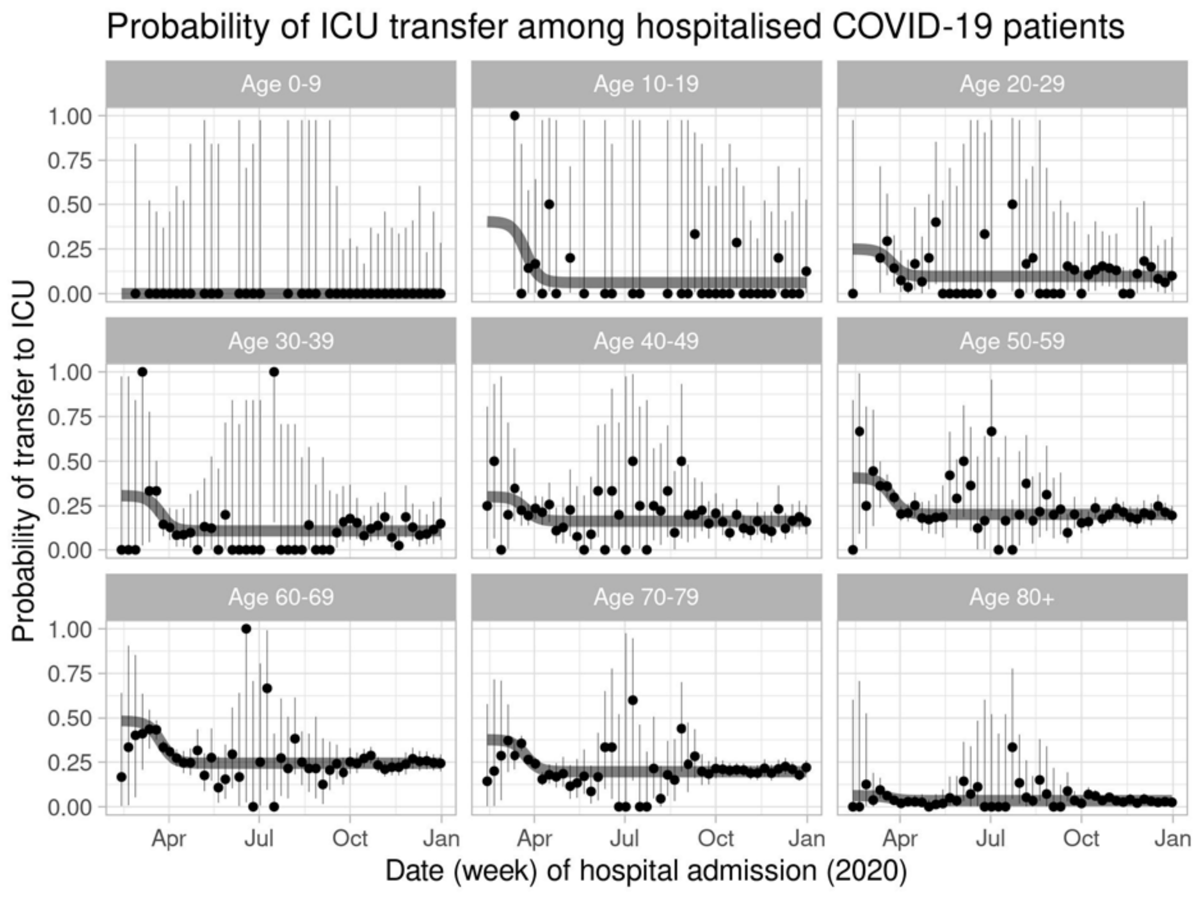


Figure S2. Estimated age-specific and time-varying probability of transfer to the ICU among patients hospitalised for COVID-19. The grey curves are the model fits. The points and vertical lines are weekly means and 95% confidence intervals.

*Clinical progression model: the probability of transfer back to a hospital ward (from ICU) (function 3)*

The probability of transfer back to a hospital ward is modelled as a logistic curve

$$\mathrm{Pr}_i^H(t) = \mathrm{Pr}_i^H(-\infty) + \left(\mathrm{Pr}_i^H(\infty) - \mathrm{Pr}_i^H(-\infty)\right)\frac{e^{r(t-t_m)}}{1+e^{r(t-t_m)}} \quad \text{(S17)}$$

The parameters are estimated by maximizing the likelihood function given the observed outcomes for all patients admitted to the ICU

$$\ell\left(\mathbf{Pr}^H(-\infty), \mathbf{Pr}^H(\infty) \middle| r, t_m, \boldsymbol{\iota}^H, \mathbf{t}, \mathbf{i}\right) = \sum_k \iota_k^H \log\left(\mathrm{Pr}_{i_k}^H(t_k)\right) + \left(1-\iota_k^H\right)\log\left(1-\mathrm{Pr}_{i_k}^H(t_k)\right) \quad \text{(S18)}$$

in which $\iota_k^H = 1$ indicates if individual $k$ has left the ICU by transfer to a hospital ward, and $\iota_k^H = 0$ if individual $k$ has left the ICU by direct discharge/death. Age groups under 40 years of age are combined because these contain only few individuals. The parameters $r$ and $t_m$ are not estimated, but fixed at the estimates obtained after maximizing Equation (S14). Because durations of stay are distributed equally for those that are transferred or are discharged/died, patients still on the ICU can be ignored. The resulting fit is shown in Figure S3.

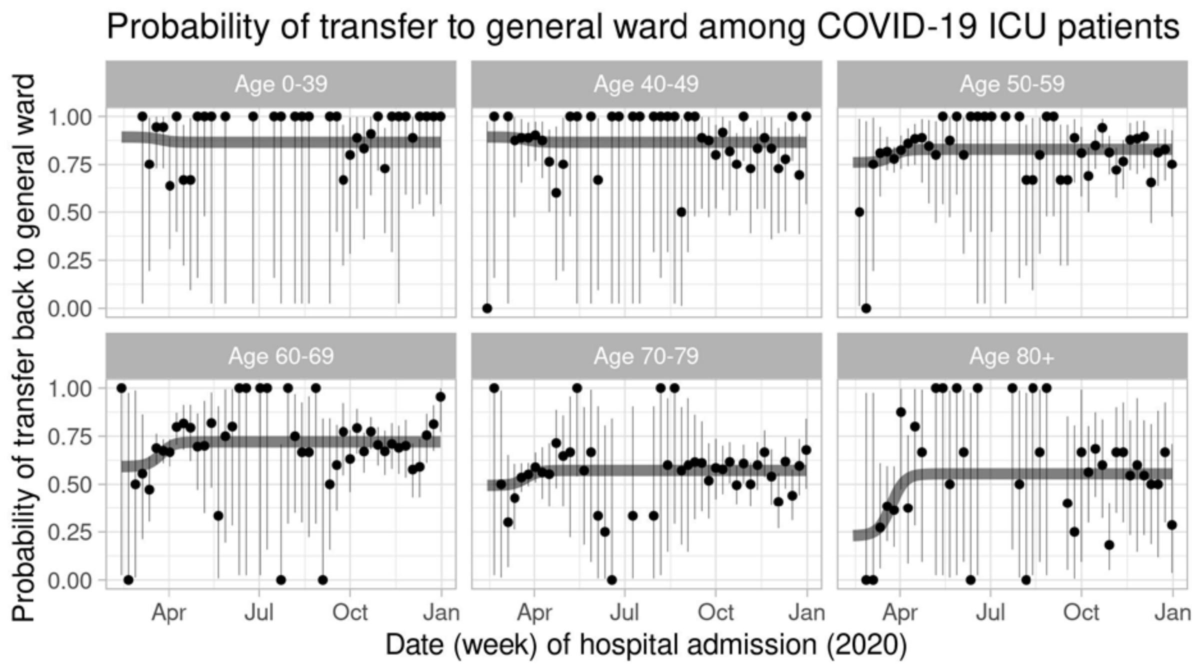


Figure S3. Estimated age-specific and time-varying probability of transfer back to the general hospital ward among patients on the ICU for COVID-19. The grey curves are the model fits. The points and vertical lines are weekly means and 95% confidence intervals.

*Clinical progression model: all delays and lengths of stay within the hospital (functions 6-9)*

The four delay and length-of-stay distributions in the hospital, from hospital admission to discharge or death (without transfer to the ICU), $d^{D_A}(t)$, from hospital admission to transfer to the ICU, $d^{ICU}(t)$, from transfer to the ICU to discharge or death, $d^{D_{ICU}}(t)$, and from re-transfer to a hospital ward to discharge or death, $d^{D_H}(t)$, are all modelled as negative binomial distributions, with time-varying means, the same for all age groups. The means $\mu^X(t)$ are modelled as sums of three logistic curves:

$$\mu^X(t) = \mu_1^X + \sum_{l=1,2,3} \left(\mu_{l+1}^X - \mu_l^X\right) \frac{e^{r_l^X\left(t-t_l^X\right)}}{1+e^{r_l^X\left(t-t_l^X\right)}} \tag{S19}$$

with time *t* being the time of admission to the hospital, for all distributions.

The parameters of these curves are estimated by maximizing the log-likelihood function given the observed delays and lengths-of-stay

$$\ell\left(\alpha^X, \boldsymbol{\mu}^X, \mathbf{t}^X, \mathbf{r}^X \middle| \mathbf{X}, \boldsymbol{\iota}^X, \mathbf{t}\right) = \sum_k \iota_k^X \log\left(p_{NB\left(\alpha^X, \mu^X(t_k)\right)}\left(X_k\right)\right) + \left(1-\iota_k^X\right)\log\left(1-P_{NB\left(\alpha^X, \mu^X(t_k)\right)}\left(X_k\right)\right) \tag{S20}$$

in which $X_k$ is the observed length-of-stay of patient *k*, $\iota_k^X = 1$ indicates if this observation is uncensored, and $\iota_k^X = 0$ if it is censored. The functions $p_{NB(\,)}(\,)$ and $P_{NB(\,)}(\,)$ are the negative binomial probability and cumulative probability, respectively. The censored observations of patients in the hospital ward that have not (yet) been transferred to the ICU are all used in the likelihood for $X = D_A$ and not for $X = ICU$. The resulting fits are shown in Figures S4, S5, S6, and S7.

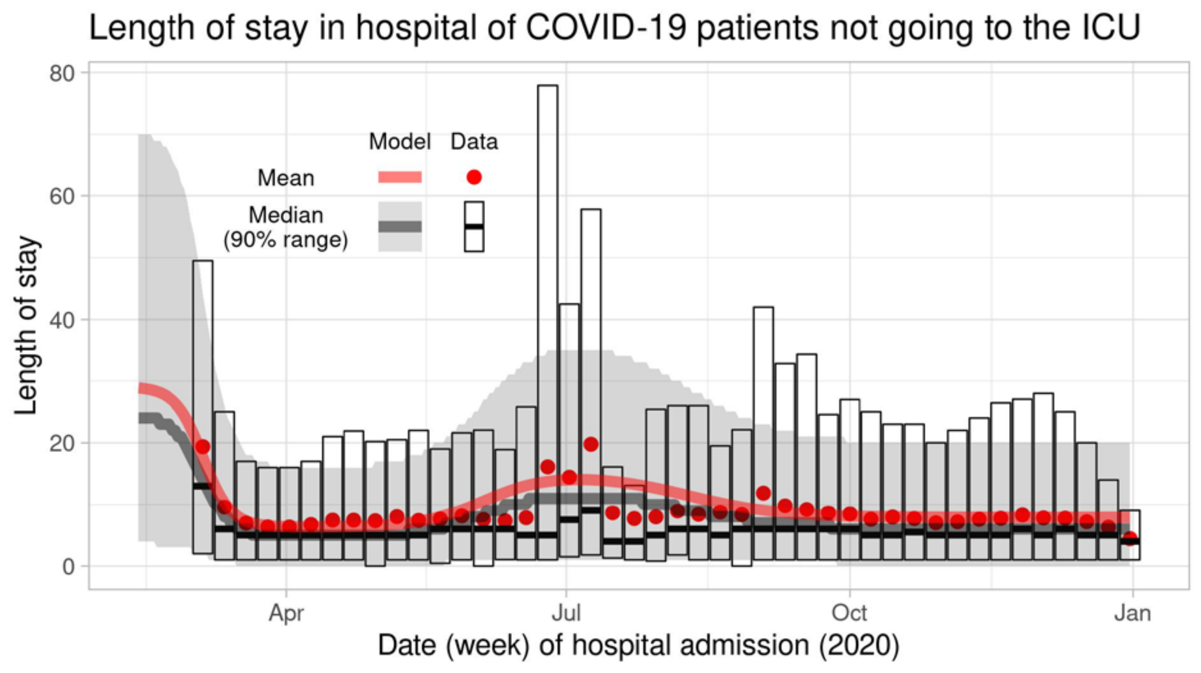


Figure S4. Time-varying distribution of the length of stay in hospital of patients not going to the ICU. The transparent lines and grey ribbon show the model fit, i.e. the negative binomial distribution used in the simulations. The points and cross-bars show the data per week, including patients still in the hospital (censored data).

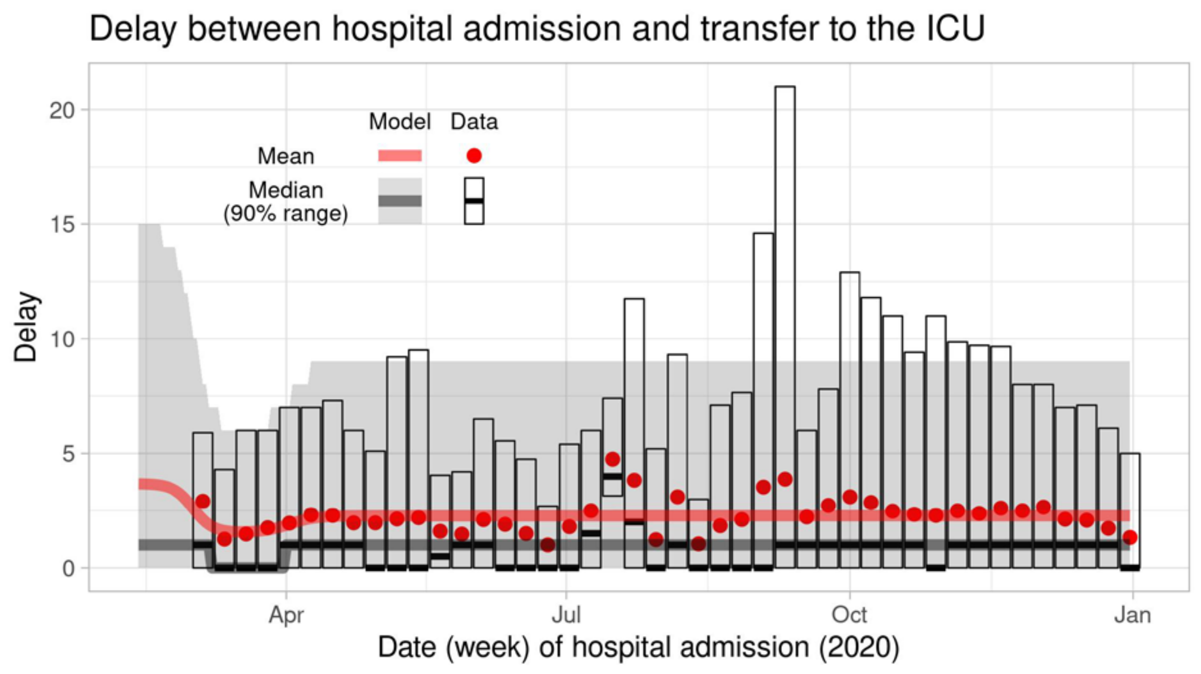


Figure S5. Time-varying distribution of the delay between hospital admission and transfer to the UCI. The transparent lines and grey ribbon show the model fit, i.e. the negative binomial distribution used in the simulations. The points and cross-bars show the data per week.

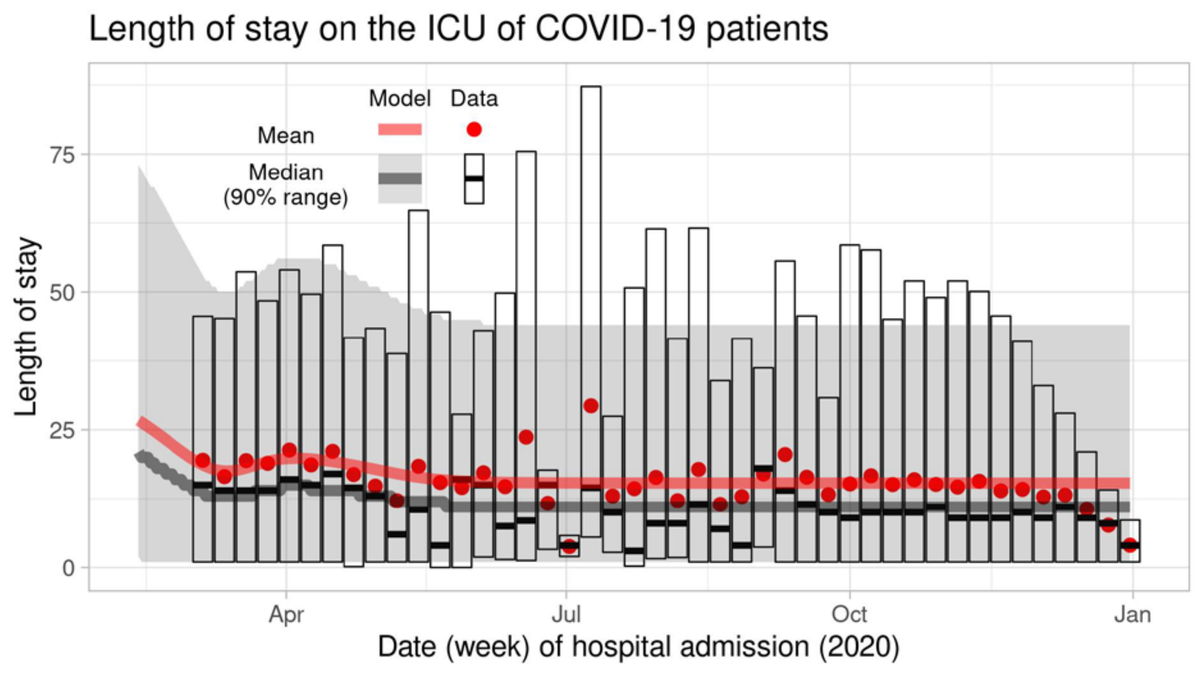


Figure S6. Time-varying distribution of the length of stay on the ICU of COVID-19 patients. The transparent lines and grey ribbon show the model fit, i.e. the negative binomial distribution used in the simulations. The points and cross-bars show the data per week, including patients still on the ICU (censored data).

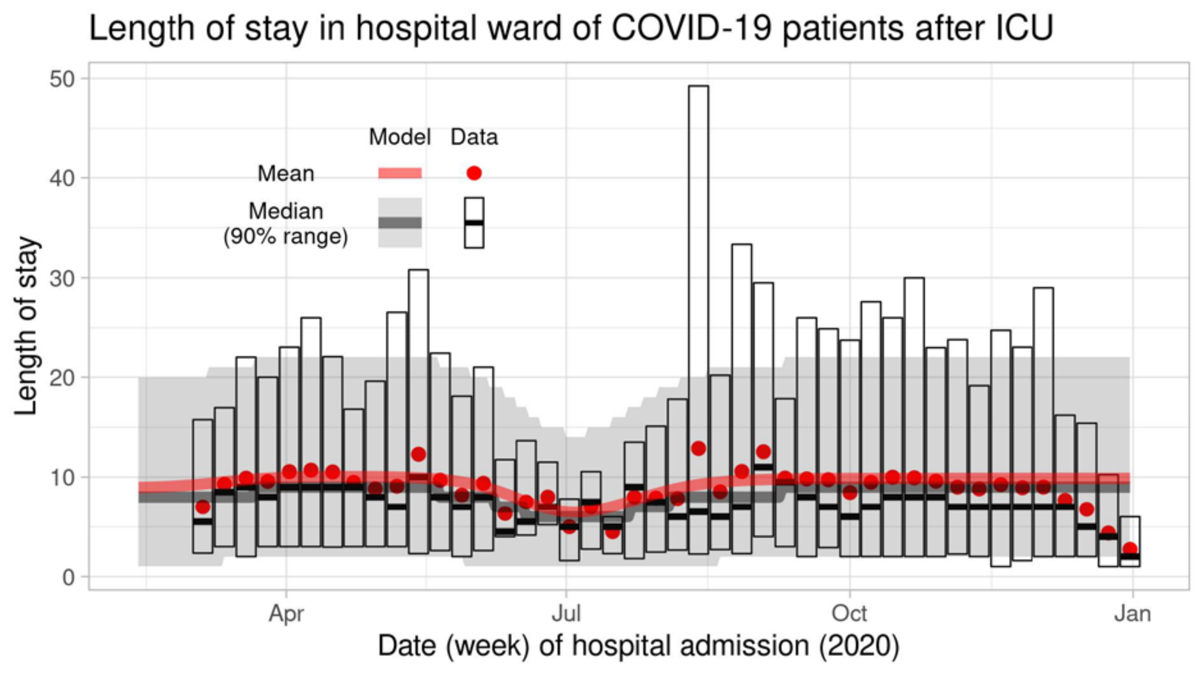


Figure S7. Time-varying distribution of the length of stay on the hospital of COVID-19 patients, after transfer from the ICU. The transparent lines and grey ribbon show the model fit, i.e. the negative binomial distribution used in the simulations. The points and cross-bars show the data per week, including patients still in the hospital (censored data).

*Observation of ICU admissions: reporting delay*

A reporting delay distribution specific for the day of the week is estimated, $p_{obs}(\tau)$, defined as the probability that a case is reported within $\tau$ days after admission to the ICU.

The data consist of admitted cases *k*, with their weekday of admission $w_k \in [0,6]$ and reporting delay $\delta_k$. For each weekday of admission, the empirical distribution of reporting delay $\Delta$ can directly be calculated from the data as

$$\Pr(\Delta \le \delta | W = w) = \frac{\sum_{k|w_k=w} 1_{\delta_k \le \delta}}{\sum_{k|w_k=w} 1} \tag{S21}$$

These seven distributions, one for each weekday, are used to construct $p_{obs}(\tau)$ for weekday $w_{obs}$, the day of analysis:

$$p_{obs}(\tau) = \Pr(\Delta \le \tau | W = (w_{obs} - \tau)\%7) \tag{S22}$$

Because the reporting delay distribution may change over time, it is estimated only with data from recent months. The distribution of 6 January 2021 is shown in Figure S8, and was estimated with data of patients admitted since 1 September 2020.

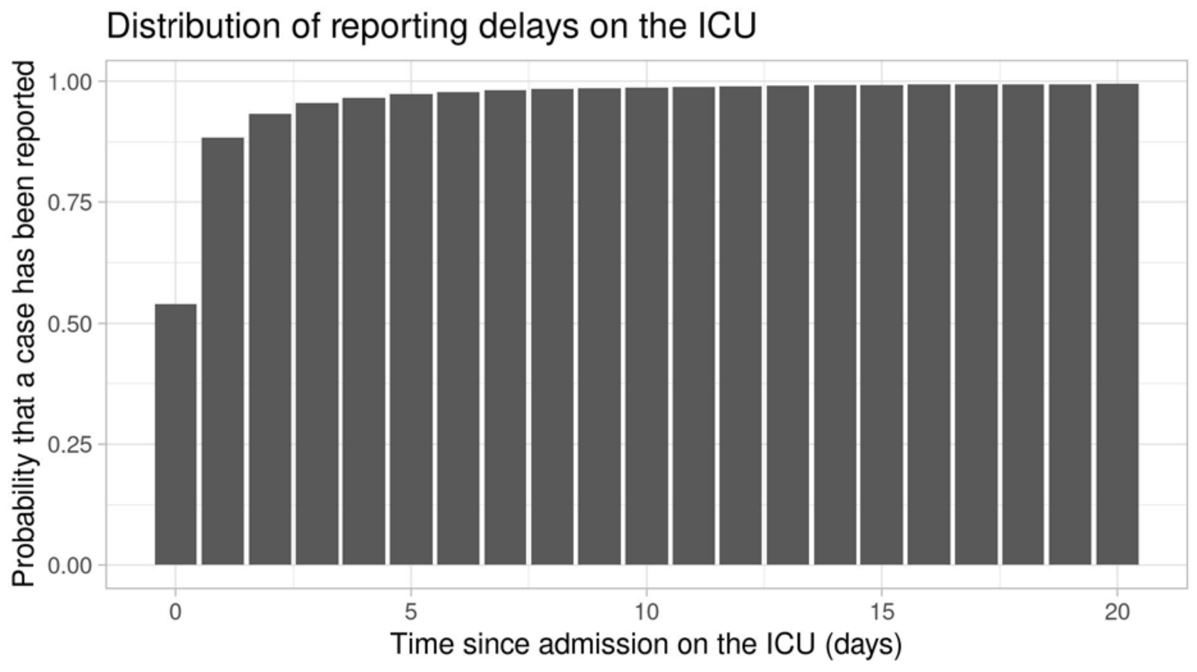


Figure S8. Reporting delay distribution used in the analysis of 6 January 2021

2.1.4. Parameter values based on notification data (OSIRIS)

The OSIRIS notification data consist of records per patient, with their age and dates of symptom onset and hospital admission. These are used to estimate the time-varying delay distribution from symptom onset to hospital admission (function 5 of the clinical progression model).

*Clinical progression model: the delay from symptom onset to hospital admission (function 5)*

The delay from symptom onset to hospital admission is modelled as a negative binomial distribution, with time-varying means specific for each age group. The means $\mu_i^A(t)$ are modelled as a logistic curve:

$$\mu_i^A(t) = \mu_{i,1}^A + \left(\mu_{i,2}^A - \mu_{i,1}^A\right)\frac{e^{r^A(t-t^A)}}{1+e^{r^A(t-t^A)}} \tag{S23}$$

The parameters of these curves are estimated by maximizing the log-likelihood

$$\ell\left(\alpha^A, \boldsymbol{\mu}_{\cdot,1}^A, \boldsymbol{\mu}_{\cdot,2}^A, r^A, t^A \middle| \mathbf{t}, \mathbf{A}\right) = \sum_k \log\left(p_{NB\left(\alpha^A, \mu_{i_k}^A(t_k)\right)}\left(A_k\right)\right) \tag{S24}$$

in which patient $k$ with age $i_k$ had a delay of $A_k$ days after symptom onset at day $t_k$ before admission to the hospital. Censored data are not available, so censoring has to be ignored. The resulting fits are shown in Figure S9.

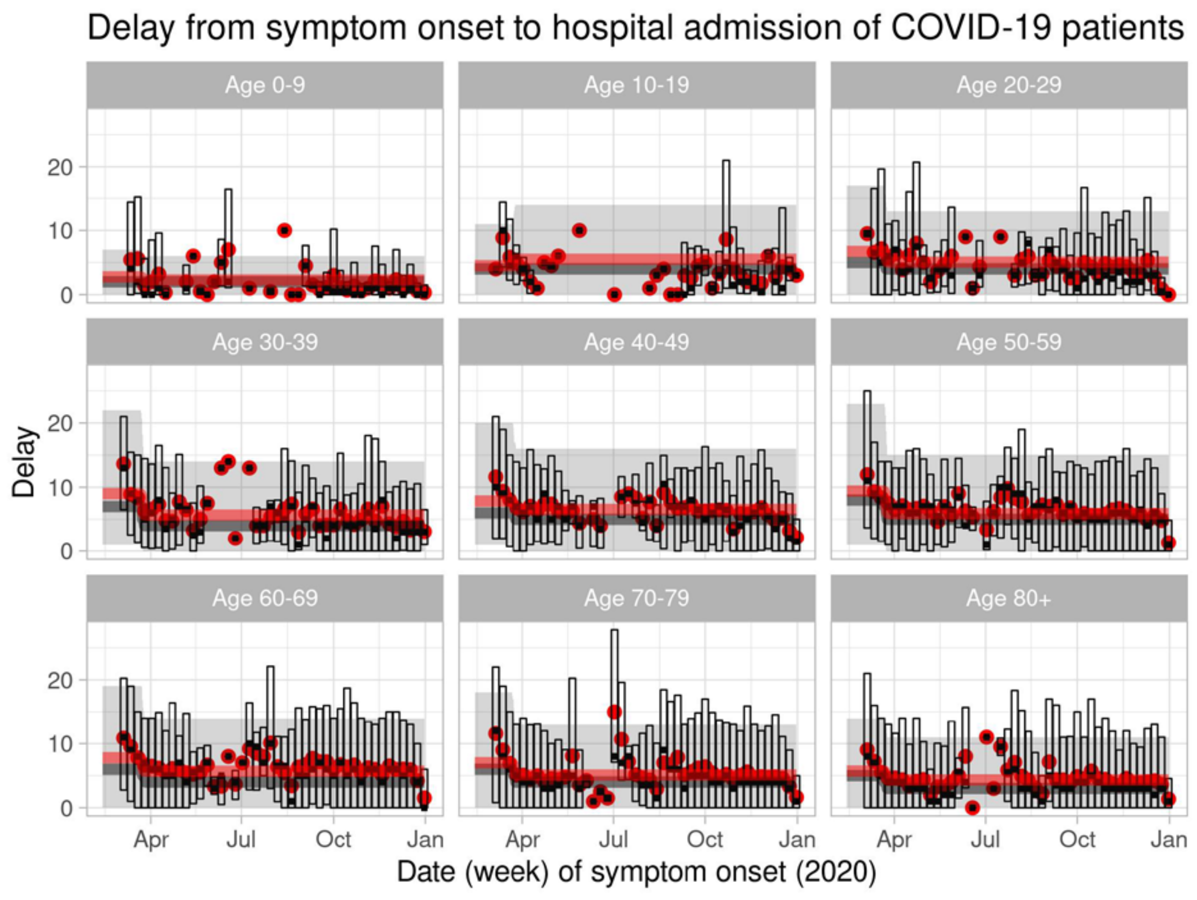


Figure S9. Time-varying and age-specific distribution of the delay between symptom onset and hospital admission of COVID-19 patients, after transfer from the ICU. The transparent lines and grey ribbon show the model fit, i.e. the negative binomial distribution used in the simulations. The points and cross-bars show the data per week. Means are shown in red, medians and 90% ranges in black and grey. For recent weeks, longer delays have not yet been observed (censored data).

2.1.5. Parameter values based serological survey PiCo

The PiCo data consist of seroprevalence estimates $p_i^{sero+}$ for each age group *i*, measured at mean survey date 15 June 2020. Assuming a mean delay from infection to seroconversion of 19 days, these seroprevalences reflect the cumulative incidence up to 27 May 2020. Incidence was very low in this period, so calculations are insensitive to this specific delay.

The PiCo data are used to estimate the age-specific susceptibility and infectivity, and to complete the estimation of the probability of hospital admission.

*Transmission model: age-specific infectivity and susceptibility*

The age-specific infectivity and susceptibility parameters $\boldsymbol{\sigma}^{\mathrm{I}}$ and $\boldsymbol{\sigma}^{\mathrm{S}}$ are effectively assumed to be equal: $\boldsymbol{\sigma}^{\mathrm{I}} = \boldsymbol{\sigma}^{\mathrm{S}} = \boldsymbol{\sigma}$. In the age-structured transmission model with next-generation matrix

$$\mathbf{M}_T(t) = \tfrac{2\beta(t)}{\gamma} \boldsymbol{\sigma}^{\mathrm{T}} \mathbf{C}_T (\boldsymbol{\sigma} \circ \mathbf{x}) \tag{S3}$$

the right eigenvector $\boldsymbol{\xi}_T(t)$ corresponding to the largest eigenvalue is equal to the age distribution of incidence of infections. For this property we can ignore all parameters that are not age-specific, so we only need the eigenvector $\boldsymbol{\xi}_T$ of

$$\mathbf{M}_T = \boldsymbol{\sigma}^{\mathrm{T}} \mathbf{C}_T (\boldsymbol{\sigma} \circ \mathbf{x}) \tag{S25}$$

The PiCo data reflect incidence up to 27 May 2020. Of all cases infected up to that day, about half were infected before lockdown and about half during lockdown (a rough approximation based on the symmetry of the epidemic incidence curve). That means that, in the model, we assume that 50% of cases were infected when contact matrix $\mathbf{C}_1$ was in place (unrestricted contacts as measured in Pienter), and 50% when contact matrix $\mathbf{C}_2$ was in place (adjusted matrix for the first phase of lockdown). Thus, we solve $\boldsymbol{\sigma}$ from

$$\boldsymbol{\xi}_1(\boldsymbol{\sigma}) + \boldsymbol{\xi}_2(\boldsymbol{\sigma}) = \mathbf{p}^{sero+} \tag{S26}$$

under the condition that

$$\sum_i \boldsymbol{\xi}_{1,i} = \sum_i \boldsymbol{\xi}_{2,i} \tag{S27}$$

to make both matrices contribute equally. The resulting vector, normalised with the lowest age group as reference group, is

$$\boldsymbol{\sigma}^{\mathrm{T}} = \begin{pmatrix} 1 & 3.05 & 5.75 & 3.54 & 3.71 & 4.36 & 5.69 & 5.32 & 7.21 \end{pmatrix}$$

This is well in line with published estimates of heterogeneity in susceptibility, e.g. Zhang et al (6) who reported that adults aged 15-64 are about three times more susceptible than children aged 0-14, and elderly aged over 65 are about 50% more susceptible than adults.

*Clinical progression model: the probability of hospital admission (function 1)*

The PICO data are used to estimate $p_i^{A+C}$ of Equation (S9). Assuming on average 12 days between infection and hospital admission and 19 days between infection and seroconversion, the seroprevalence on 15 June 2020 (the mean survey date) reflects the cumulative incidence of infections of 27 May 2020, and the cumulative number of hospital admissions of 8 June 2020.

From the PiCo data with seroprevalence $p_i^{sero+}$ in age group *i*, the cumulative number of infections $Y_i^{PiCo}$ in age group *i* on 27 May 2020 is estimated as

$$Y_i^{PiCo} = Nx_i p_i^{sero+} \tag{S28}$$

Using Equation (S9), from the NICE data with daily hospitalisations $A_i(t)$ in age group *i*, the cumulative number of infections $Y_i^{NICE}$ on 27 May 2020 is estimated as

$$Y_i^{NICE} = \sum_{t \le 8\,June\,2020} A_i(t) / \Pr_i^A(t) = \frac{1}{p_i^{A+C}} \sum_{t \le 8\,June\,2020} A_i(t)\left(1 - p_i^{D|A}(t)\right) \tag{S29}$$

These two estimators are used to estimate $p_i^{A+C}$ by solving $Y_i^{PiCo} = Y_i^{NICE}$, with $p_i^{D|A}(t)$ as estimated from the NICE data (Equation (S14)). With this result, the probability of admission (Equation (S9)) can be calculated (Figure S10).

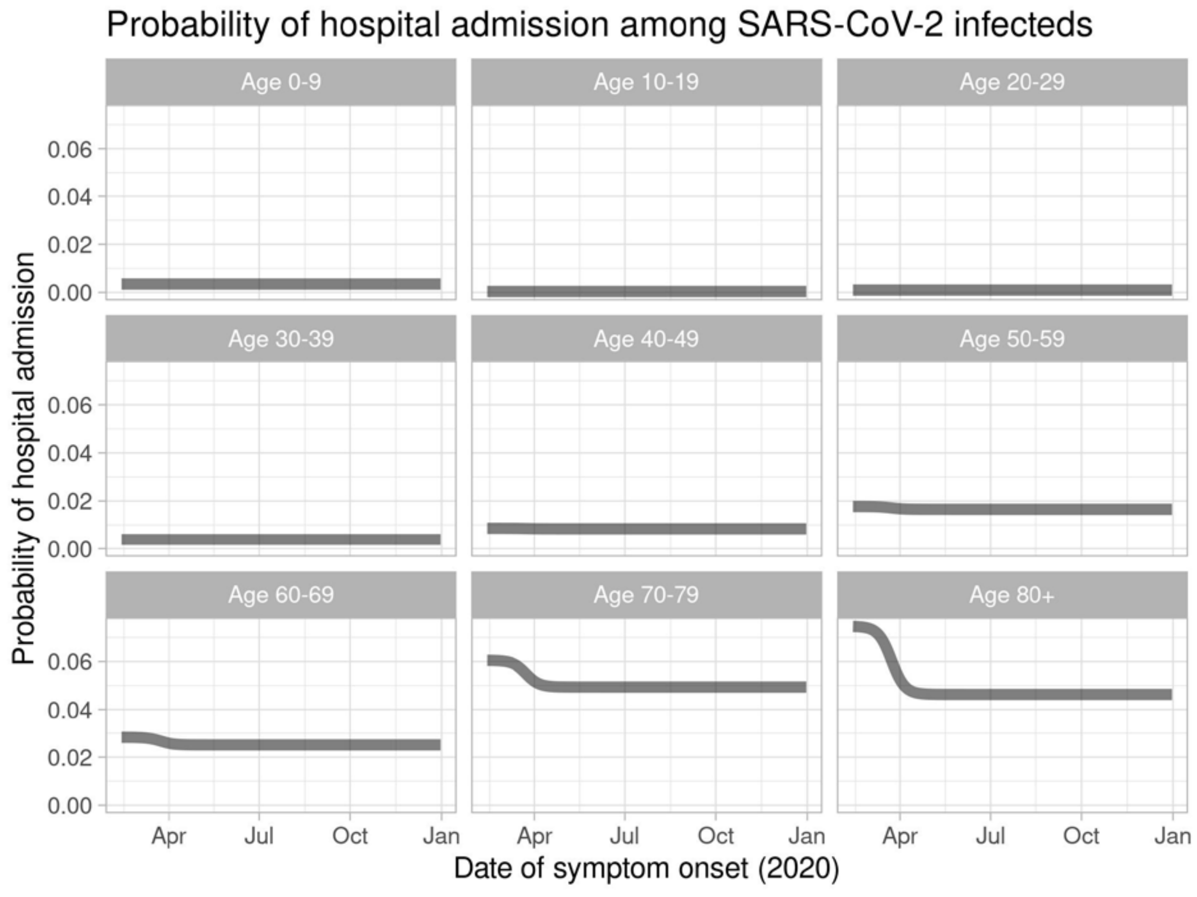


Figure S10. Estimated age-specific and time-varying probability of hospital admission

2.2. Step 2: parameter estimation for age-specific contact patterns

In the model, contact matrices $\mathbf{C}_T$ describe how different age groups interact with each other in consecutive periods $T$ of the pandemic during which different sets of control measures were in place. Each matrix is used up to transition time $u_T$, the time of policy change. An exception is made for the rapid sequential deployment of control measures mid-March 2020, with associated adaptations in contact behaviour. For this period we assume the existence of two transition times, with the same contact matrix after the first and second transition. The dates of these transitions, $u_1$ and $u_2$, are estimated in step 3 (below); note that these dates do not necessarily correspond to the actual days when major policy changes were implemented.

The matrices are based on contact data collected for the Pienter study in 2017, before COVID-19 (7). In this contact survey, participants recorded their contacts over the course of a day in different settings: home, school, work, leisure, transport, and rest. COVID-19 measures can affect these contacts; how much, in which setting and in which age group will depend on the specific measure. Estimating the reduction in contacts, something that cannot be measured, is done by treating it as a Fermi problem, i.e. by breaking it down into many smaller estimation problems which reduces the expected error of the problem as a whole (8, 9). For each set of COVID-19 measures, two researchers independently predict how the measures will affect the contacts of each age group in each setting, while ensuring consistency with previous sets of measures. This yields the Fermi estimates, i.e. consensus estimates for the relative contact rates with an uncertainty range.

In the simulations of 6 January 2021, twelve different matrices are used for in fourteen periods $T$ (the same matrices were used in periods 1 and 2, and in periods 10 and 12). These twelve matrices describe different sets of control measures in place during different periods of the first year of the epidemic. Table S3 at the bottom of the Supplement gives an overview of the estimated relative contact rates in each of the six contact settings (home, work, school, transport, leisure, other), during each of those periods. The keywords refer to the control measures that were in place.

To make 200 matrices for each set of control measures, relative contact rates were uniformly sampled from intervals with indicated values as midpoint, and widths of 0.2. With these sampled reductions, the contact frequencies in the original Pienter contact data are changed (using the age of the participant), and contact matrices are estimated as described in Van de Kassteele (10), which ensures reciprocity of contact rates. Thus, 200 contact matrices are obtained to describe the effect of a given set of COVID-19 control measures. On 28 May 2020, new estimates were made for all sets of control measures, to match the observations of the first CoMix contact study (11, 12).

#### 2.3. Step 3: estimation of the transmissibility parameter

The transmissibility parameters $\beta_T$ and matrix transition times $u_T$ are required for the rate of transmission $\beta(t)$. Together with the initial number of infected persons $y_0$ on 12 February 2020, all $\beta_T$, $u_1$, and $u_2$ are estimated by fitting the model to the ICU admission data, conditional on all parameter values as inferred in steps 1 and 2, and all transition time $u_T$ for $T > 2$. We use the optim function in R version 3.6.0 (13) to maximize the log-likelihood, with $T_{\max}$ stepwise constant transmissibilities in $\beta(t)$:

$$\ell\left(y(0),u_1,u_2,\beta_1,\ldots,\beta_{T_{\max}}\middle|Z(\ ),u_3,\ldots,u_{T_{\max}},\theta,\mathbf{C}_1,\ldots,\mathbf{C}_{T_{\max}}\right)=$$
$$\sum_t Z(t)\log\left(p_{obs}\left(t_{analysis}-t\right)\sum_i z_i\left(t;y(0),\beta_1,\ldots,\beta_{T_{\max}},\theta,u_1,\ldots u_{T_{\max}},\mathbf{C}_1,\ldots,\mathbf{C}_{T_{\max}}\right)\right) \quad \text{(S30)}$$

Here, $Z(\ )$ are the daily Poisson-distributed observed ICU admissions, $\theta$ all parameters estimated in step 1, $\mathbf{C}_T$ the means of all sets of 200 sampled contact matrices, $p_{obs}\left(t_{analysis}-t\right)$ the probability of reporting at the day of analysis, and $z_i(\ )$ the simulated number of admissions.

At each later transition time $u_T$, the change in contact matrices from $C_T$ to $C_{T+1}$ should reflect the anticipated changes in contact behaviour caused by the policy change at that moment. If this is indeed the case, it can be assumed that $\beta_{T+1}=\beta_T$, thus reducing the number of model parameters. As a consequence, the stepwise constant transmissibility parameter $\beta(t)$ will have fewer changepoints (where $\beta_{T+1}\neq\beta_T$) than there are transition times. To select changepoints, We fitted models with changepoints at different subsets of transition times $u_T$, and compared these fits with Akaike's Information Criterion (AIC, (14)). Each week, we selected the set of changepoints with the lowest value of the AIC. From the final selected model, point estimates are obtained with a covariance matrix for all stepwise constant transmissibilities and the initial number of infected persons, calculated from the Hessian matrix obtained with the optim function in R. The resulting estimates and confidence intervals are given in section 3.1 of this Supplement, with a correlation matrix calculated from the covariance matrix.

In practice, the changepoint selection did not mean that we tried every combination of changepoints every week; that would have taken too much time. Instead, we started with the changepoints from the week before, and tried other sets by removing changepoints, moving changepoints forwards and backwards in time (to other transition times), and adding changepoints. In most weeks, this was done by starting with the most recent transition times, and when there was evidence that changepoints should be changed, earlier transition times were included in that process. Sometimes, when we made changes to the model or data analysis, we did a more extensive search along the whole time series, which sometimes also led to new estimates for the transition times $u_1$ and $u_2$. The evolution of all selected changepoints and estimates for $u_1$ and $u_2$ is shown in section 3.2 below.

#### 3. Supplemental analyses and results

##### 3.1. Complete results of analysis step 3, and comparison of the ODE and discrete-time models

Tables S1 and S2 give the results of estimation step 3 for the data of 6 January 2021, i.e. the estimated initial state $y_0$ and transmissibility parameters $\beta_T$ with 95% confidence intervals in Table S1, and the correlation matrix of these estimates in Table S2. It turns out that most confidence intervals are narrow, and that the uncertainty is highly correlated between initial state and transmissibility at the start of the pandemic, and between each subsequent pair of transmissibility parameters. It should be noted that $\beta_5$ (15 Oct – 25 Oct) was assumed to be equal to $\beta_3$ (29 Mar – 29 Sep). That is because, at the time, we thought that the two weeks between these periods (30 Sep – 14 Oct) could have been a short interruption of the already six-months continuous $\beta_3$. In line with the our general workflow as explained in the main text (section 2.5), we stayed with this decision because new changepoint analyses never rejected it.

As of 25 November 2020 we replaced the continuous-time model by the discrete-time model. In the transition phase, we carefully checked that both models gave similar results. To show their similarity, we fitted the data of 6 January 2021 to the continuous-time model and to the discrete-time model. On 6 January 2021, there were seven parameters to be estimated in Step 3: initial incidence $y_0$ and six transmissibility parameters $\beta_T$ for the periods as shown in Figure 2a (main text). Table S1 below shows the parameter estimates for the two models, Table S2 the correlation matrices of the estimates, and Figure S11 shows the fitted ICU admission curves, and their difference.

It turns out that the estimated transmissibilities are almost identical, as are the correlation matrices. The initial state values $y_0$ are different, because the initial state is parameterized differently. In the discrete-time model, $y_0/12$ is equal to a constant incidence from 1 February 2020 – 12 February 2020 (equal in each age class). In the continuous-time model, $y_0/4$ is the initial value for all states $E_1$, $E_2$, $I_1$, and $I_2$ (equal in each age class).

Table S1. Parameter estimates on the log-scale (95% CI) of Step 3 (6 January 2021), with the discrete-time and continuous-time models.

| Model parameter | Discrete-time model | Continuous-time model |
|---|---|---|
| Initial state $\log(y_0)$ [a] | 1.71 (1.29 ; 2.12) | 3.48 (3.08 ; 3.88) |
| $\log(\beta_1)$ (until 18 Mar 2020) | 5.23 (5.19 ; 5.27) | 5.22 (5.18 ; 5.26) |
| $\log(\beta_2)$ (from 19 Mar to 28 Mar 2020) | 4.71 (4.64 ; 4.79) | 4.72 (4.65 ; 4.79) |
| $\log(\beta_3)$ (from 29 Mar to 28 Sep 2020) [b] | 5.20 (5.19 ; 5.20) | 5.20 (5.19 ; 5.20) |
| $\log(\beta_4)$ (from 29 Sep to 14 Oct 2020) | 5.37 (5.34 ; 5.40) | 5.37 (5.34 ; 5.39) |
| $\log(\beta_5)$ (from 15 Oct to 25 Oct 2020) [b] | 5.20 (5.19 ; 5.20) | 5.20 (5.19 ; 5.20) |
| $\log(\beta_6)$ (from 26 Oct to 18 Nov 2020) | 5.27 (5.25 ; 5.30) | 5.27 (5.25 ; 5.30) |
| $\log(\beta_7)$ (from 19 Nov onwards) | 5.51 (5.49 ; 5.53) | 5.51 (5.50 ; 5.53) |

[a] the initial state has a different parameterisation in the two models, and can therefore not be compared

[b] a single transmissibility parameter was estimated for periods 3 and 5

Table S2. Correlation matrix of estimates of initial state and transmissibility parameters, with the discrete-time and continuous-time models[a]

Discrete-time model

| | $\log(y_0)$ | $\log(\beta_1)$ | $\log(\beta_2)$ | $\log(\beta_3)$ | $\log(\beta_4)$ | $\log(\beta_6)$ | $\log(\beta_7)$ |
|---|---|---|---|---|---|---|---|
| $\log(y_0)$ | 1 | -0.991 | 0.484 | -0.150 | 0.038 | -0.001 | -0.001 |
| $\log(\beta_1)$ | -0.991 | 1 | -0.567 | 0.246 | -0.039 | 0.010 | 0.014 |
| $\log(\beta_2)$ | 0.484 | -0.567 | 1 | -0.583 | 0.174 | 0.003 | 0.010 |
| $\log(\beta_3)$ | -0.150 | 0.246 | -0.583 | 1 | -0.554 | 0.212 | 0.098 |
| $\log(\beta_4)$ | 0.038 | -0.039 | 0.174 | -0.554 | 1 | -0.494 | 0.187 |
| $\log(\beta_6)$ | -0.001 | 0.010 | 0.003 | 0.212 | -0.494 | 1 | -0.756 |
| $\log(\beta_7)$ | -0.001 | 0.014 | 0.010 | 0.098 | 0.187 | -0.756 | 1 |

Continuous-time model

| | $\log(y_0)$ | $\log(\beta_1)$ | $\log(\beta_2)$ | $\log(\beta_3)$ | $\log(\beta_4)$ | $\log(\beta_6)$ | $\log(\beta_7)$ |
|---|---|---|---|---|---|---|---|
| $\log(y_0)$ | 1 | -0.990 | 0.478 | -0.142 | 0.039 | 0.000 | -0.001 |
| $\log(\beta_1)$ | -0.990 | 1 | -0.564 | 0.242 | -0.040 | 0.010 | 0.014 |
| $\log(\beta_2)$ | 0.478 | -0.564 | 1 | -0.577 | 0.173 | 0.004 | 0.010 |
| $\log(\beta_3)$ | -0.142 | 0.242 | -0.577 | 1 | -0.554 | 0.213 | 0.099 |

| | | | | | | | |
|---|---|---|---|---|---|---|---|
| $\log(\beta_4)$ | 0.039 | -0.040 | 0.173 | -0.554 | 1 | -0.494 | 0.187 |
| $\log(\beta_6)$ | 0.000 | 0.010 | 0.004 | 0.213 | -0.494 | 1 | -0.756 |
| $\log(\beta_7)$ | -0.001 | 0.014 | 0.010 | 0.099 | 0.187 | -0.756 | 1 |

[a] $\log(\beta_5)$ is missing, because a single transmissibility parameter was estimated for periods 3 and 5

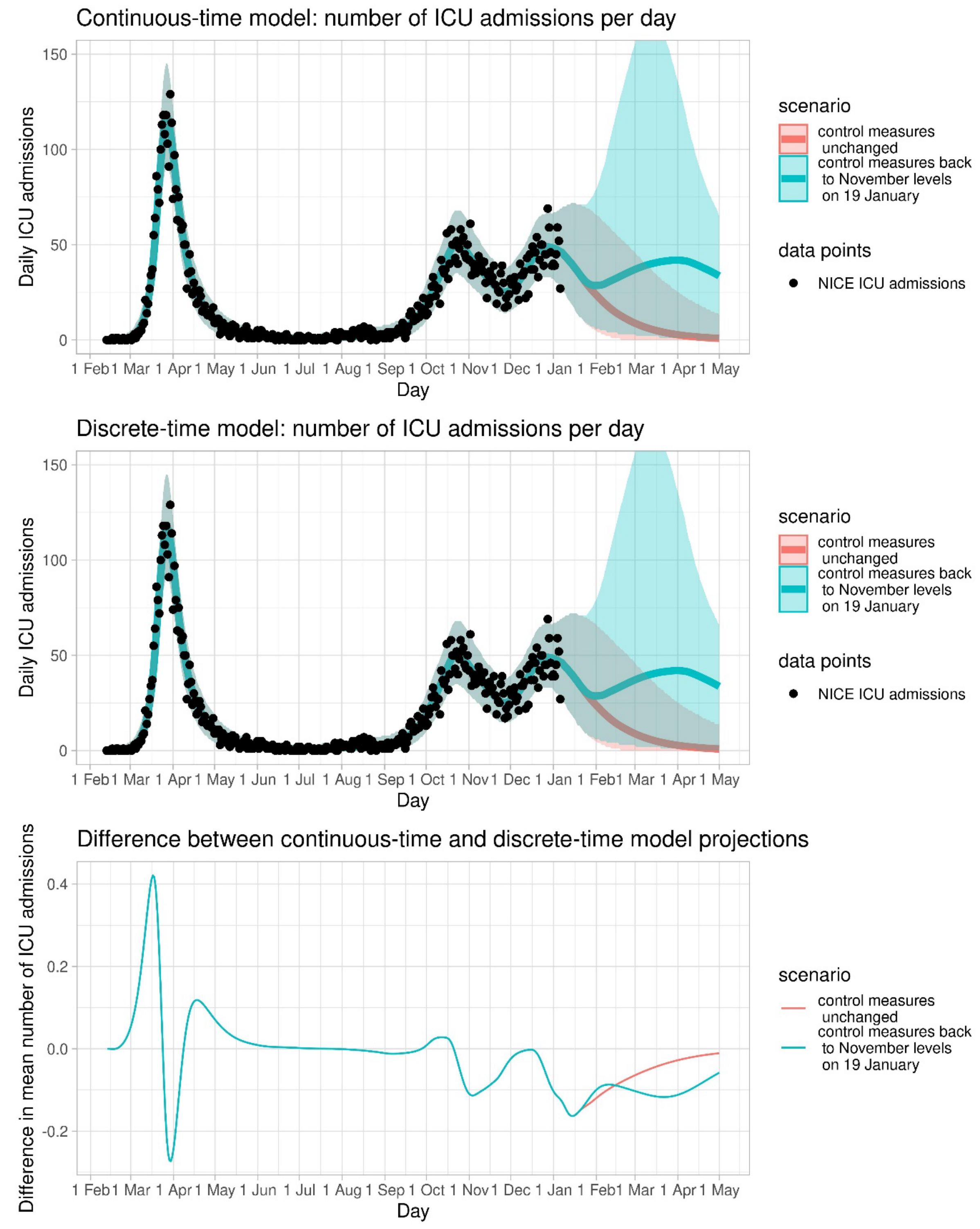


Figure S11. Model projections with a projection window of 4 months to illustrate that the continuous-time and discrete-time models produce almost identical results, also beyond the data to which the models were fitted, and also when incidence increases in the future.

3.2. Weekly evolution of transmissiblity parameter estimates

Each week the model was fitted to the most recent ICU admission data in Step 3 of the parameter estimation. This resulted in new estimates of transmissibility parameters $\beta_T$ and a selection of changepoints. To illustrate the evolution of the parameter estimates and selected changepoints, we fitted the model of 6 January 2021 to reconstructed datasets for the days $t_{analysis}$ of the original 40 model fits, with transition days $u_1$ and $u_2$ as selected in those analyses. Datasets were reconstructed by taking the dataset of observed daily ICU admissions on 6 January 2021 $Z(t)$ up to $t_{analysis}$, multiplying the observations by $p_{obs}(t_{analysis} - t)$ and rounding to the nearest integer, to simulate the reporting delay. The reason for not showing the original analyses is that the model itself has also gradually changed, as well as the data for Step 1. Now all that is different between the 40 fits is $t_{analysis}$, and the transition days $u_1$ and $u_2$.

Figure S12 shows the estimated values for $\log(\beta_T)$ and the identified changepoints, when the colours change from left to right. It is clearly visible how incoming data adjust identification of changepoints and estimates of transmissibility, so that periods in which model projections were incorrect (March, July, November) were corrected in the weeks thereafter. Also, the transition days $u_1$ and $u_2$ are sometimes adjusted, also long after they occurred; all these changes had to do with changes in the clinical progression model, which had consequences for simulation of the entire ICU admission time series, or with changes in interpreting the NICE hospital data (re-admissions occurring towards the end of 2020 made it necessary to redefine admission and duration of stay).

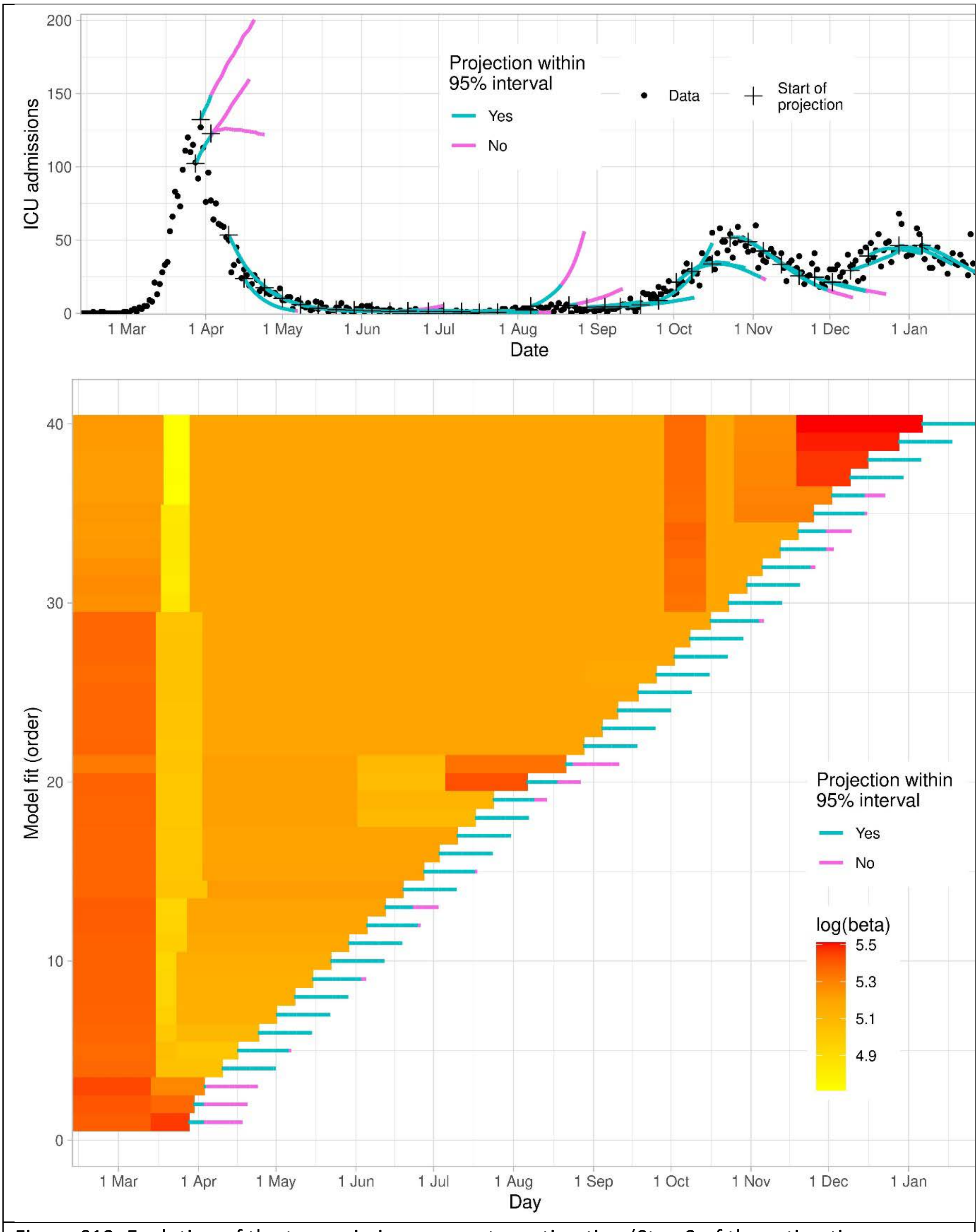


Figure S12. Evolution of the transmission parameter estimation (Step 3 of the estimation procedure). Top panel: all 40 projections of ICU admissions, with colours indicating whether the realized admissions were within the 95% interval. Bottom panel: transmission parameters log(beta) for all 40 model fits. Each row is one model fit, carried out at the day at which the orange-red bar ends. Each bar is followed by a projection line, matching the projections in the top panel. Changes in log(beta) in a row indicate identified changepoints.

Table S3: Control periods *T* with transition times $u_T$, and Fermi estimates of relative contact rates, per set of control measures, contact setting, and age group. The estimates were used as midpoints of intervals of width 0.2, from which 200 uniform samples were taken to obtain 200 contact matrices per set of control measures.

| Keyword description of (changes in) control measures, and time period in place | Contact setting | Age group | Fermi estimate of relative contact rate (relative to Pienter contact survey of 2017) |
|---|---|---|---|
| Pienter 2017 (from start to $u_1$ = 18 Mar 2020) | All | 0-100 | 1.00 |
| Calibrated on CoMix (from 19 Mar to $u_2$ = 28 Mar, and from 29 Mar to $u_3$ = 10 May 2020)<br>Lockdown: schools closed, work from home, keeping distance outside, leisure closed. Shops were allowed to open, but most shops closed | **Name: Contactmatrices_intelligentlockdown_march** | | |
| | Home | 0-100 | 1.00 |
| | Work | 0-100 | 0.483 |
| | School | 0-100 | 0.214 |
| | Transport | 0-100 | 0.344 |
| | Leisure | 0-100 | 0.102 |
| | Other | 0-100 | 0.889 |
| Batch 1 (from 11 May to $u_4$ = 1 June 2020)<br>Primary schools and day care open at 50%<br>Sports for under 12s<br>Contact professions<br>Leisure allowed outside<br>Shops, markets, libraries | **Name: Contactmatrices_batch1_11may** | | |
| | Home | 0-100 | 1.00 |
| | Work | 0-100 | 0.53 |
| | School | 0-10<br>10-20<br>20-100 | 0.61<br>0.32<br>0.214 |
| | Transport | 0-100 | 0.39 |
| | Leisure | 0-10<br>10-20<br>20-100 | 0.30<br>0.16<br>0.102 |
| | Other | 0-100 | 0.90 |
| Batch 2 (from 2 June to $u_5$ = 5 July 2020)<br>Primary schools and day care full open<br>Secondary schools<br>Terraces<br>Leisure inside < 30p<br>Museums<br>Facemask in public transport | **Name: Contactmatrices_batch2_1june** | | |
| | Home | 0-100 | 1.00 |
| | Work | 0-100 | 0.63 |
| | School | 0-10<br>10-20<br>20-100 | 1.00<br>0.64<br>0.214 |
| | Transport | 0-100 | 0.53 |
| | Leisure | 0-10<br>10-20<br>20-100 | 0.45<br>0.42<br>0.30 |
| | Other | 0-100 | 0.95 |
| Batch 3 (from 6 July to $u_6$ = 30 Aug 2020)<br>Leisure inside < 100p or no max<br>Events < 100p<br>Campings<br>Sports inside | **Name: Contactmatrices_batch3_summerholiday** | | |
| | Home | 0-100 | 1.00 |
| | Work | 0-100 | 0.63 |
| | School | 0-100 | 0 |
| | Transport | 0-100 | 0.64 |
| | Leisure | 0-20<br>20-100 | 0.80<br>0.60 |
| | Other | 0-100 | 1.00 |
| Start school year (from 31 Aug to $u_7$ = 28 Sep 2020)<br>Schools fully open; universities not<br>More work contacts | **Name: Contactmatrices_start-schoolyear-september** | | |
| | Home | 0-100 | 1.00 |
| | Work | 0-100 | 0.77 |
| | School | 0-10<br>10-20<br>20-100 | 1.00<br>0.84<br>0.214 |

| | | | |
|---|---|---|---|
| | Transport | 0-100 | 0.74 |
| | Leisure | 0-40<br>40-100 | 0.80<br>0.60 |
| | Other | 0-100 | 1.00 |
| Autumn restrictions (from 29 Sep to $u_8$ = 14 Oct 2020)<br>Schools fully open; universities not<br>Working from home advised; bars close at 22:00, max group sizes inside and outside; max 3 people at home | **Name: Contactmatrices_restrictions28sep** | | |
| | Home | 0-100 | 0.90 |
| | Work | 0-100 | 0.69 |
| | School | 0-10<br>10-20<br>20-100 | 1.00<br>0.84<br>0.214 |
| | Transport | 0-100 | 0.58 |
| | Leisure | 0-20<br>20-100 | 0.60<br>0.30 |
| | Other | 0-100 | 0.95 |
| Extreme autumn restrictions (from 15 Oct to $u_9$ = 25 Oct 2020) with holiday<br>Schools half open; universities not<br>Working from home required; bars closed, max group sizes inside and outside further reduced; max 3 people at home | **Name: Contactmatrices_partiallockdown-october-holiday** | | |
| | Home | 0-100 | 0.90 |
| | Work | 0-100 | 0.57 |
| | School | 0-10<br>10-20<br>20-100 | 0.61<br>0.53<br>0.214 |
| | Transport | 0-100 | 0.46 |
| | Leisure | 0-10<br>10-20<br>20-100 | 0.60<br>0.40<br>0.20 |
| | Other | 0-100 | 0.95 |
| Extreme autumn restrictions (from 26 Oct to $u_{10}$ = 4 Nov 2020 & from 19 Nov to $u_{12}$ = 14 Dec 2020)<br>Schools fully open; universities not<br>Working from home required; bars closed, max group sizes inside and outside further reduced; max 3 people at home | **Name: Contactmatrices_partiallockdown-october** | | |
| | Home | 0-100 | 0.90 |
| | Work | 0-100 | 0.60 |
| | School | 0-10<br>10-20<br>20-100 | 1.00<br>0.84<br>0.214 |
| | Transport | 0-100 | 0.48 |
| | Leisure | 0-10<br>10-20<br>20-100 | 0.60<br>0.40<br>0.20 |
| | Other | 0-100 | 0.95 |
| 2 weeks november extra restrictions (from 5 Nov to $u_{11}$ = 18 nov 2020)<br>Schools fully open; universities not<br>Working from home required; bars closed, all leisure closed; sport clubs open but no groups; max 2 people outside; max 2 people invited at home | **Name: Contactmatrices_2week-lockdown-november** | | |
| | Home | 0-100 | 0.85 |
| | Work | 0-100 | 0.58 |
| | School | 0-10<br>10-20<br>20-100 | 1.00<br>0.84<br>0.214 |
| | Transport | 0-100 | 0.48 |
| | Leisure | 0-10<br>10-20<br>20-100 | 0.40<br>0.25<br>0.15 |
| | Other | 0-100 | 0.95 |
| Winter lockdown (from 15 Dec to $u_{13}$ = 20 Dec 2020 & from 4 Jan to 22 Jan 2021)<br>Schools, universities closed; working from home required; non-essential shops closed, bars closed, all leisure closed; | **Name: Contactmatrices_winter-lockdown** | | |
| | Home | 0-100 | 0.85 |
| | Work | 0-100 | 0.483 |
| | School | 0-100 | 0.214 |
| | Transport | 0-100 | 0.36 |
| | Leisure | 0-10<br>10-20 | 0.30<br>0.20 |

| | | | |
|---|---|---|---|
| sport clubs only open outside for under 18's; max 2 people outside; max 2 people invited at home | | 20-100 | 0.102 |
| | Other | 0-100 | 0.889 |
| Christmas holiday with winter lockdown (from 21 Dec 2020 to $u_{14}$ = 3 Jan 2021)<br>Schools, universities closed; working from home required; non-essential shops closed, bars closed, all leisure closed; sport clubs only open outside for under 18's; max 2 people outside; max 2 people invited at home (3 during christmas days) | **Name: Contactmatrices_winter-lockdown-christmas** | | |
| | Home | 0-100 | 1.00 |
| | Work | 0-100 | 0.43 |
| | School | 0-100 | 0.214 |
| | Transport | 0-100 | 0.33 |
| | Leisure | 0-10<br>10-20<br>20-100 | 0.30<br>0.20<br>0.102 |
| | Other | 0-100 | 0.95 |


1. Diekmann O, Heesterbeek H, Britton T. Mathematical tools for understanding infectious disease dynamics. Princeton, N.J.: Princeton University Press; 2013. xiv, 502 pages p.
2. Ganyani T, Kremer C, Chen D, Torneri A, Faes C, Wallinga J, Hens N. Estimating the generation interval for coronavirus disease (COVID-19) based on symptom onset data, March 2020. Euro Surveill. 2020;25(17).
3. Tindale LC, Stockdale JE, Coombe M, Garlock ES, Lau WYV, Saraswat M, et al. Evidence for transmission of COVID-19 prior to symptom onset. Elife. 2020;9.
4. Backer JA, Klinkenberg D, Wallinga J. Incubation period of 2019 novel coronavirus (2019-nCoV) infections among travellers from Wuhan, China, 20-28 January 2020. Euro Surveill. 2020;25(5).
5. Lauer SA, Grantz KH, Bi Q, Jones FK, Zheng Q, Meredith HR, et al. The Incubation Period of Coronavirus Disease 2019 (COVID-19) From Publicly Reported Confirmed Cases: Estimation and Application. Ann Intern Med. 2020;172(9):577-82.
6. Zhang J, Litvinova M, Liang Y, Wang Y, Wang W, Zhao S, et al. Changes in contact patterns shape the dynamics of the COVID-19 outbreak in China. Science. 2020;368(6498):1481-6.
7. Verberk JDM, Vos RA, Mollema L, van Vliet J, van Weert JWM, de Melker HE, van der Klis FRM. Third national biobank for population-based seroprevalence studies in the Netherlands, including the Caribbean Netherlands. BMC Infect Dis. 2019;19(1):470.
8. Bergstrom CT, West JD. Calling bullshit : the art of skepticism in a data-driven world. First edition. ed. New York: Random House; 2020. xvi, 318 pages p.
9. Wikipedia. Fermi Problem: Wikipedia; [Available from: https://en.wikipedia.org/wiki/Fermi_problem.
10. Van de Kassteele J, Van Eijkeren, J., Wallinga, J. Efficient estimation of age-specific social contact rates between men and women. The Annals of Applied Statistics. 2017;11(1):320-39.
11. Backer JA, Bogaardt, L., Beutels, P., Coletti, P., Edmunds, W.J., Gimma, A., Van Hagen, C.E.C., Hens, N., Jarvis, C.I., Vos, E.R.A., Wambua, J., Wong, D., Van Zandvoort, K., Wallinga, J. Dynamics of non-household contacts during the COVID-19 pandemic in 2020 and 2021 in the Netherlands. Sci Rep. 2023;13:5166.
12. Verelst F, Hermans L, Vercruysse S, Gimma A, Coletti P, Backer JA, et al. SOCRATES-CoMix: a platform for timely and open-source contact mixing data during and in between COVID-19 surges and interventions in over 20 European countries. BMC Med. 2021;19(1):254.

13. Team RC. R: A language and environment for statistical computing. Vienna, Austria.: R foundation for statistical computing; 2019.
14. Burnham KPA, D.R. Model selection and multimodel inference. A practical information-theoretic approach. 2nd ed. New York: Springer Science + Business Media; 2002.
